# Socioeconomic Position and Falls Among Middle- and Older-Aged Adults: A Systematic Review with a Life Course Approach

**DOI:** 10.1101/2025.05.13.25327493

**Authors:** Frerik Smit, Marcia R Franco, Catherine Sherrington, Stéphane Cullati, Fiona Blyth, Anita van Zwieten, Saman Khalatbari-Soltani

## Abstract

**Background:** Falls among middle- and older-aged adults are a significant public health concern. However, a holistic understanding of how different indicators of socioeconomic position (SEP) are associated with falls is lacking, particularly for SEP across the life course.

**Methods:** We systematically searched for observational studies analysing the association between at least one indicator of SEP and one fall outcome. Due to heterogeneity between included studies, results were narratively synthesised.

**Results:** After de-duplication, 5,880 search results were screened and 125 studies were included. Only 14 included studies explicitly aimed to study the relationship between SEP and falls, which generally found that higher SEP was associated with lower risks/rates of falls. An additional nine studies also had relevant adjusted models that also largely showed a protective relationship. However, adjusted results were mixed and often lacked statistical significance. The remaining 102 studies only contained unadjusted results of interest, with 50%-100% of results for each SEP indicator showing that low SEP groups experience disproportionately high risks/rates of fall outcomes compared to high SEP groups. Notably, only four studies measured any SEP indicators from a stage of the life course prior to the study period.

**Conclusions:** Our findings suggest that falls disproportionately impact low SEP groups and that knowledge gaps exist regarding the relationship between different SEP indicators and falls, particularly for SEP exposures across the life course. Future research on this topic should utilise causal diagrams for appropriate model building and include a wide range of SEP indicators across the life course.

## Introduction

Falls, defined as “inadvertently coming to rest on the ground, floor or lower level” (1), are a significant public health issue common among middle-aged adults (2) that increases rapidly with age (1), with 26.5% of older people falling at least once yearly (3). Falls can lead to severe injuries and death – representing the second highest cause of unintentional injury deaths worldwide (4). With an ageing population and projected increases in fall rates (5), preventing falls is of increasing urgency.

It is well established that individual- and neighbourhood-level socioeconomic position (SEP) significantly impacts various health outcomes (6), including healthy ageing and frailty (7,8). However, how SEP across the life course impacts falls remains unclear. SEP refers to the resource- and prestige-based factors that contribute to the position individuals or groups hold within societal structures and is commonly determined by an aggregate of social and economic factors, including education, income, and occupation (9). Life course conceptual models that consider the timing (e.g., sensitive and critical period models), duration (e.g., accumulation model), and sequencing (e.g., pathway and social mobility models) of exposure to socioeconomic (dis)advantages may help explain how SEP across the life course impacts falls in later stages of life. For instance, the sensitive period model suggests that the impact of an exposure on health outcomes in later life is greater when that exposure occurs in certain sensitive periods. The strict accumulation model suggests that health outcomes are determined by the cumulative effect of the frequency and duration of exposures rather than the timing of exposures. The pathway model emphasises the sequential ordering of exposures, and the social mobility model focuses on the trajectories of exposures between different life course periods (10,11). To date, no review has considered a life course perspective on this topic. Further, we are only aware of one existing review that studied the relationship between SEP indicators and falls, but it focused exclusively on community-dwelling older adults (12).

We aimed to address these knowledge gaps by systematically reviewing existing literature on 1) the association between SEP at any stage of the life course and falls among middle- and older-aged adults, and 2) life course approaches and conceptual models used in studies of associations between SEP and falls.

## Methods

This systematic review followed the Preferred Reporting Items for Systematic Reviews and Meta-Analyses (PRISMA) 2020 checklist (appendix p2) (13). It also followed a protocol registered in the International Prospective Registration of Systematic Reviews (PROSPERO) (http://www.crd.york.ac.uk/PROSPERO) (CRD42024534813) and published elsewhere (14).

### Search Strategy and Selection Criteria

Searches were conducted from inception to March 15^th^ 2024 in MEDLINE, Embase, and PsycINFO via Ovid using MeSH headings and search terms related to SEP, falls, middle-/older-aged adults, and observational studies (appendix p4).

Eligible studies were observational studies (cross-sectional, longitudinal cohort, and case-control designs) published in peer-reviewed journals, comprising of participants with a mean or median age of > 40 years that reported results on the epidemiological/aetiological association between a SEP indicator at any stage of the life course and a fall outcome. SEP indicators of interest included education, income, housing, wealth, occupation, employment, and social status (9). Fall outcomes included fall occurrences, recurrent falls, (non)injurious falls, fall-related hospitalizations, and fall deaths. Studies with a risk factor or predictive modelling design were only eligible if they reported unadjusted or minimally adjusted results (i.e., models adjusted for a limited number of pre-defined covariates such as age and sex). This was done in order to avoid overinterpreting findings from non-aetiological studies given potential issues with confounding, overadjustment bias, and Table 2 fallacy (15–18). To elaborate, overadjustment bias refers to analytical adjustment (e.g., covariate adjustment in a regression model) that introduces bias rather than removing bias (18). This bias is commonly found in social epidemiological studies and generally arises due to intermediate and collider variables being naively and inappropriately treated as confounders (18). Table 2 fallacy is a related phenomenon, whereby variables are mutually adjusted for in a multivariable model, and their estimates are uniformly interpreted even though the only estimate that reflects its aetiological association with the outcome variable is the primary exposure variable of interest (17). Qualitative and ecological studies, reviews, commentaries, case series/studies, dissertations/theses, books, reports, working papers, and conference abstracts were excluded. No restrictions were placed on geography, language, year of study publication, and study population (except for age).

### Study Selection

Using Covidence software (http://covidence.org), search results were de-duplicated, and titles and abstracts were screened by one reviewer, with 20% screened independently by a second reviewer (94.9% inter-rater agreement). Full texts were subsequently screened by two reviewers independently. Disagreements were resolved through inter-reviewer discussions and third-reviewer consultation.

### Data Extraction

One reviewer extracted data into a standardised Microsoft Excel data extraction form which was cross-checked by a second reviewer. Data related to study and participant characteristics, SEP exposures and fall outcomes, study results, and life-course considerations. For studies specifically analysing the association between measures of SEP and falls, we extracted both unadjusted and adjusted results, while for risk factor, predictive modelling, and other studies not explicitly focused on SEP and falls, we only extracted relevant minimally-adjusted and/or unadjusted results.

### Risk of Bias Assessment

A modified Quality in Prognosis Studies (QUIPS) (19) critical appraisal tool (appendix pp5-8) was used independently by two reviewers to assess the risk of bias (ROB) of studies explicitly aiming to study SEP and falls and other studies with relevant adjusted results. The tool was modified by changing the term “prognostic factor” to “exposure variable” and including additional items on overadjustment bias (20,21) – an important source of bias in health inequities studies (18) – from the Risk of Bias in Non-Randomised Studies of Exposure (ROBINS-E) tool (22). ROB assessments were not conducted on the studies not explicitly aiming to study SEP and falls from which we only extracted unadjusted results, as they would all be categorised as having a high risk of bias simply by nature of their study type. Moreover, given that these studies had alternative aims than studying SEP and falls, appraising their quality with that of the other included studies was not deemed appropriate.

### Data Synthesis

Using an evidence-based mapping technique (23), considerable heterogeneity between studies was found (appendix pp9-13), thus we narratively synthesised results. Results in the lowest and highest SEP categories were compared, consistent with prior systematic reviews (7,24). Results were classified as indicating either a protective (i.e., higher SEP associated with a lower risk/rate of falls), adverse (i.e., higher SEP associated with a higher risk/rate of falls), mixed, or no association (i.e., null estimate of effect (25)) between SEP and falls. To avoid direct comparisons between studies with different statistical analysis approaches, we reported the results of the three different groups of studies separately (studies explicitly aiming to study SEP and falls, other studies with relevant adjusted models, and other studies only containing relevant unadjusted results). The main reported synthesis focused on the most appropriate model (confounder adjusted models with the least amount of mediators) (21), determined according to simplified causal diagrams (appendix p14) developed for this review. The results of additional models are reported in the appendix (pp41-44). For studies without any adjusted models of interest, we report unadjusted results in terms of the relative distribution of falls by SEP indicator due to their high risk of confounding bias (26). Finally, we also report a synthesis of studies that adopted a life course approach to SEP and falls.

## Results

Our search yielded 7,152 results, from which 125 studies were included (Figure 1; appendix p15). All results are stratified into studies explicitly aiming to study SEP and falls (n=14), studies not explicitly aiming to study SEP and falls with adjusted models of interest (n=9), and studies not explicitly aiming to study SEP and falls with only unadjusted results of interest (n=102).

**Figure 1.**
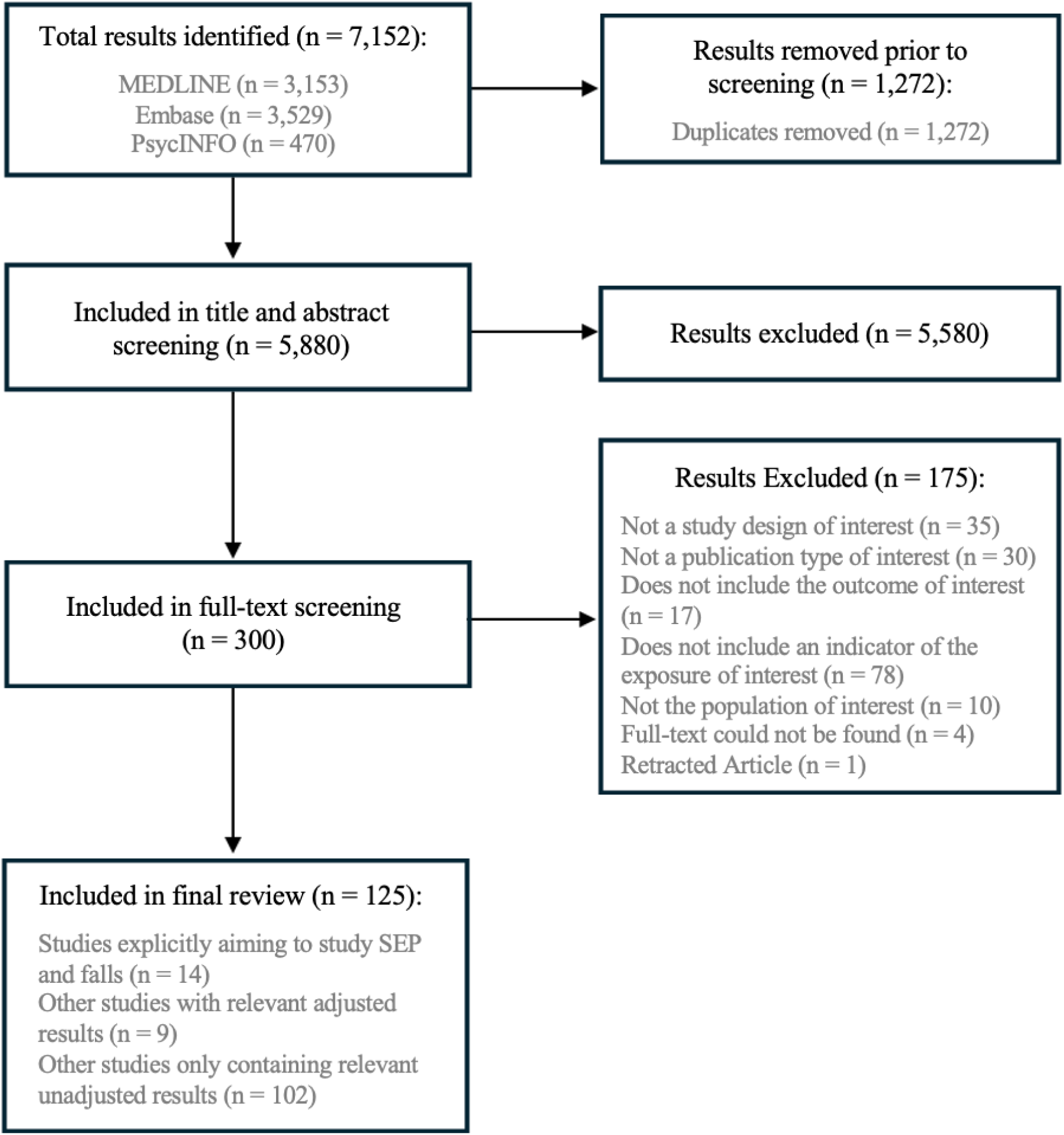
**PRISMA flow chart of search and study selection process**

Across included studies, most were cross-sectional (61%), published after 2010 (84%) and based in the Americas (35%), Western Pacific (24%), or Europe (22%) (Table 1). Most studies also included less than 10,000 participants (85%), had majority older adults 60+ (82%) and majority female (78%). The overall characteristics of the 14 studies focused on SEP and falls are similar to the entire sample, except that none were based in Africa, Southeast-Asia, or the Eastern Mediterranean. Further detailed characteristics of these studies can be found in Table 2 and in the appendix (pp16-36). Specific considerations, including number of socioeconomic indicators and overadjustment bias are reported in the appendix (p37). Detailed results of the ROB assessments are reported and synthesised in the appendix (pp38-40). In summary, they showed that a high risk of bias was common for overadjustment (41%), outcome measurement (39%), and study attrition (36%).

**Table 1.** Summary overview of study characteristics.

|  | All included studies<br>(n=125)<br>n (%) | Studies focused on SEP and falls (n=14)<br>n (%) | Other studies with relevant adjusted models (n=9)<br>n (%) | Other studies with only unadjusted results of interest (n=102)<br>n (%) |
| --- | --- | --- | --- | --- |
| **Study type** |  |  |  |  |
| Cross-sectional | 76 (60.8%) | 7 (50%) | 3 (33%) | 66 (64.7%) |
| Longitudinal | 41 (32.8%) | 7 (50%) | 4 (44%) | 30 (29.4%) |
| Case-control | 8 (6.4%) | .. | 2 (22%) | 6 (5.9%) |
| **Publication date** |  |  |  |  |
| Before 2000 | 3 (2.4%) | .. | 2 (22%) | 1 (1.0%) |
| 2000-2009 | 17 (13.6%) | 3 (21%) | 2 (22%) | 12 (11.8%) |
| 2010 onwards | 105 (84.0%) | 11 (79%) | 5 (56%) | 89 (87.3%) |
| **Geographic location (WHO region)** |  |  |  |  |
| African Region | 6 (4.8%) | .. | .. | 6 (5.9%) |
| Region of the Americas | 44 (35.2%) | 6 (43%) | 5 (56%) | 33 (32.4%) |
| South-East Asian Region | 15 (12.0%) | .. | .. | 15 (14.7%) |
| European Region | 27 (21.6%) | 5 (36%) | 2 (22%) | 20 (19.6%) |
| Eastern Mediterranean Region | 8 (6.4%) | .. | .. | 8 (7.8%) |
| Western Pacific Region | 30 (24.0%) | 3 (21%) | 2 (22%) | 25 (24.5%) |
| **Sample size** |  |  |  |  |
| <1,000 | 53 (42.4%) | 2 (14%) | 4 (44%) | 47 (46.1%) |
| 1,000 to <10,000 | 53 (42.4%) | 8 (57%) | 4 (44%) | 41 (40.2%) |
| 10,000+ | 19 (15.2%) | 4 (29%) | 1 (11%) | 14 (13.7%) |
| **Mean or median age (years)** |  |  |  |  |
| >40 to <60 | 18 (14.4%) | 2 (14%) | 2 (22%) | 14 (13.7%) |
| 60+ | 102 (81.6%) | 11 (79%) | 7 (78%) | 84 (82.4%) |
| Unclear | 5 (4.0%) | 1 (7%) | .. | 4 (3.9%) |
| **Distribution of sex (% female)** |  |  |  |  |
| 0% to <25% | 7 (5.6%) | 2 (14%) | 2 (22%) | 3 (2.9%) |
| 25% to <50% | 14 (11.2%) | 1 (7%) | .. | 13 (12.7%) |
| 50% to <75% | 85 (68.0%) | 10 (71%) | 5 (56%) | 70 (68.6%) |
| 75%+ | 12 (9.6%) | .. | .. | 12 (11.8%) |
| Unclear | 7 (5.6%) | 1 (7%) | 2 (22%) | 4 (3.9%) |
| **Fall outcomes\*** |  |  |  |  |
| Fall occurrences | 98 (78.4%) | 10 (71%) | 6 (67%) | 82 (80.4%) |
| Injurious falls | 20 (16.0%) | 2 (14%) | 3 (33%) | 15 (14.7%) |
| Recurrent falls | 17 (13.6%) | 2 (14%) | .. | 15 (14.7%) |
| Fall deaths | 2 (1.6%) | 1 (7%) | .. | 1 (1.0%) |
| Inpatient falls | 3 (2.4%) | .. | .. | 3 (2.9%) |
| Non-injurious falls | 1 (0.8%) | .. | .. | 1 (1.0%) |
| **SEP measures studied\*** |  |  |  |  |
| Education | 100 (80.0%) | 10 (71%) † | 6 (67%) | 84 (82.4%) |
| Income | 36 (28.8%) | 6 (43%) † | 2 (22%) | 28 (27.5%) |
| Employment | 15 (12.0%) | .. | 1 (11%) | 14 (13.7%) |
| Occupation | 11 (8.8%) | 4 (29%) | 1 (11%) | 6 (5.9%) |
| Housing | 8 (6.4%) | 3 (21%) † | .. | 5 (4.9%) |
| Wealth | 3 (2.4%) | .. | .. | 3 (2.9%) |
| Other individual-/household-level indicator | 26 (20.8%) | 8 (57%) † | .. | 18 (17.6%) |
| Neighbourhood-/area- level indicator | 7 (5.6%) | 3 (21%) | 1 (11%) | 3 (2.9%) |
| **Life course approaches** |  |  |  |  |
| Timing | 4 (3.2%) | 3 (21%) | 1 (11%) | .. |
| Duration | 1 (0.8%) | 1 (7%) | .. | .. |
| Sequencing | .. | .. | .. | .. |

Abbreviations: SEP, socioeconomic position; WHO, World Health Organization.
\* adds up to more than 100% as studies could report multiple exposures and/or outcomes
† includes both adjusted and unadjusted results (education: 5 unadjusted, income: 1 unadjusted, housing: 2 unadjusted, other individual-/household-level indicator: 3 unadjusted)

**Table 2.**
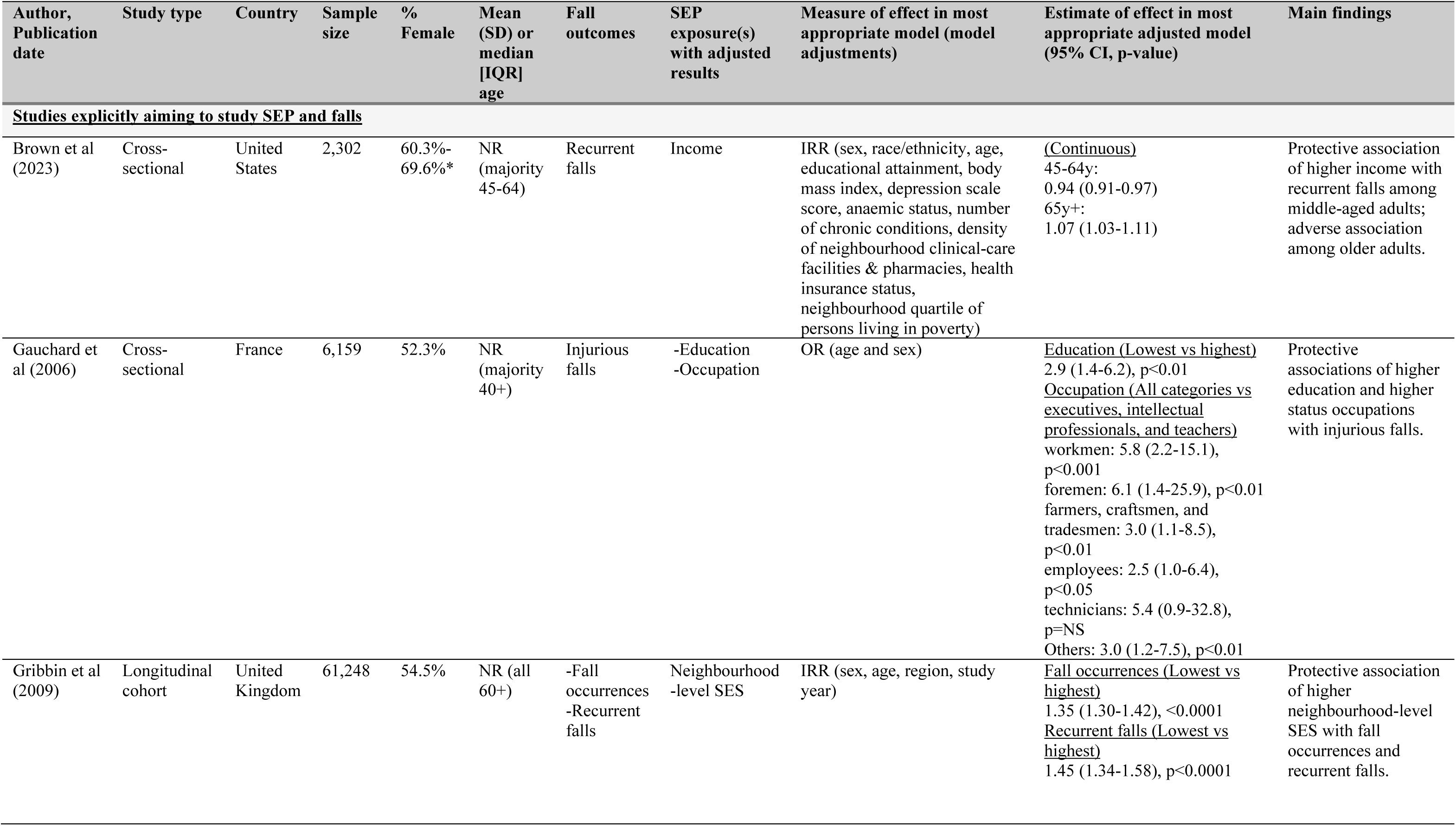
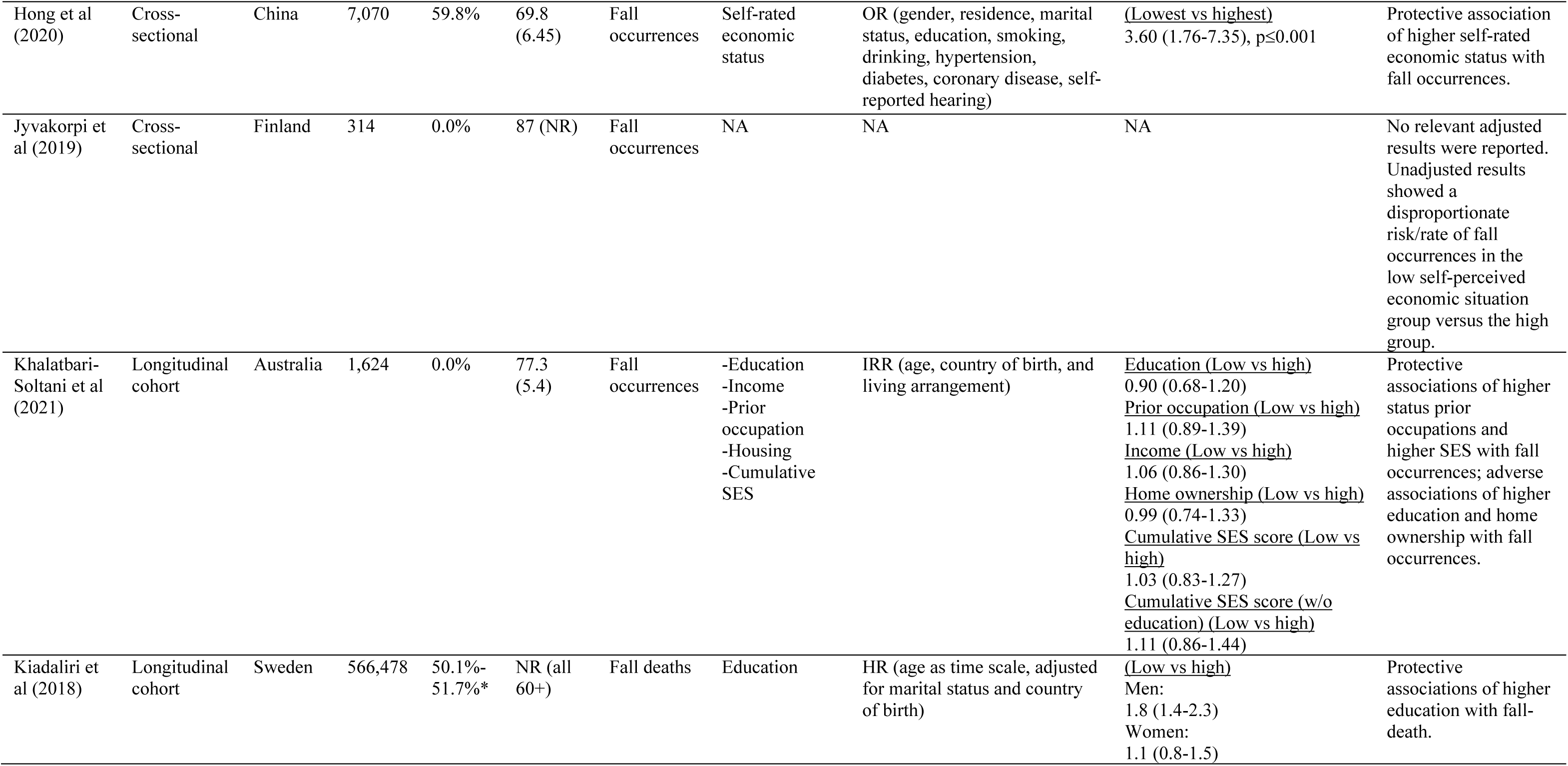
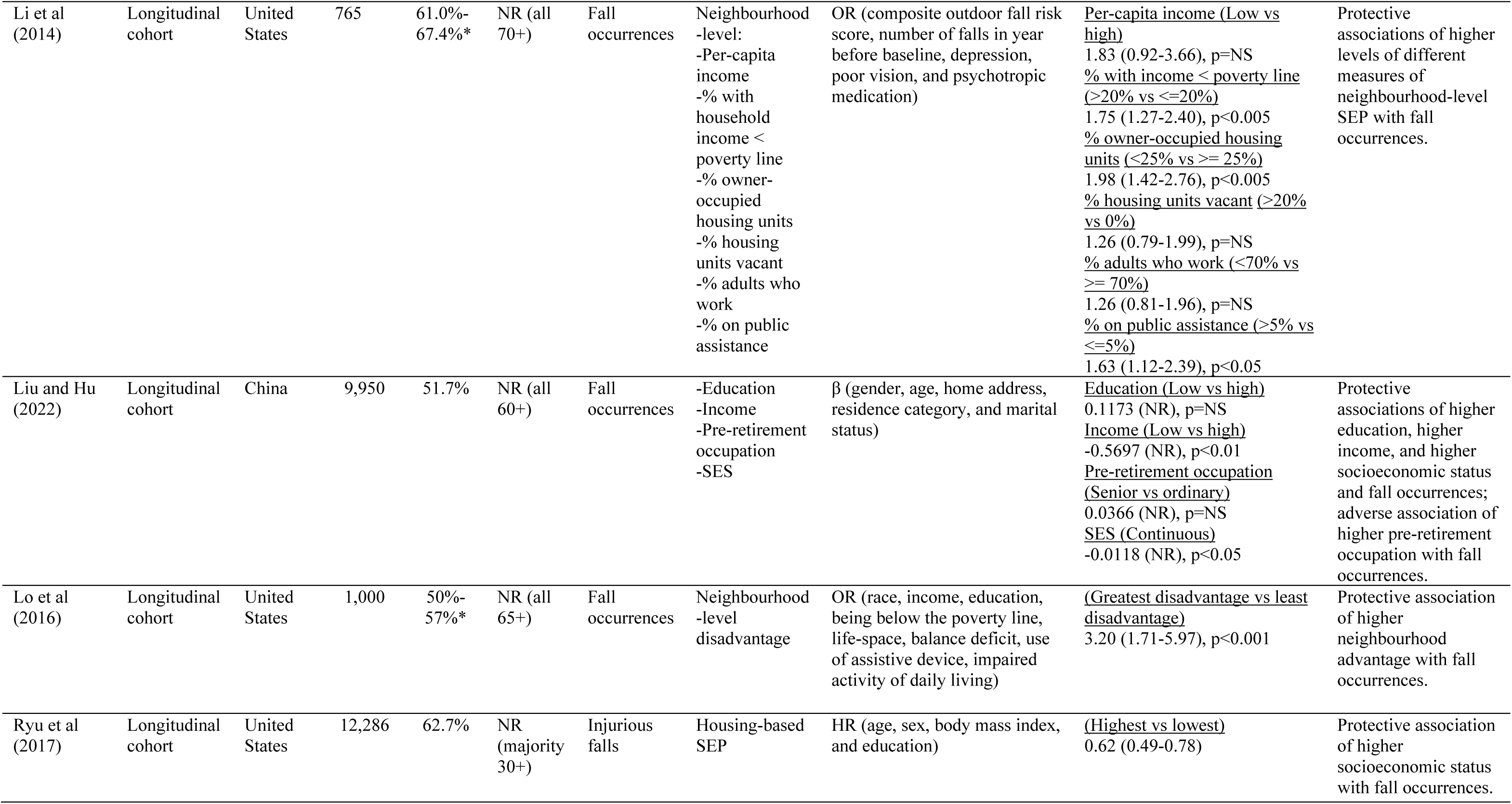
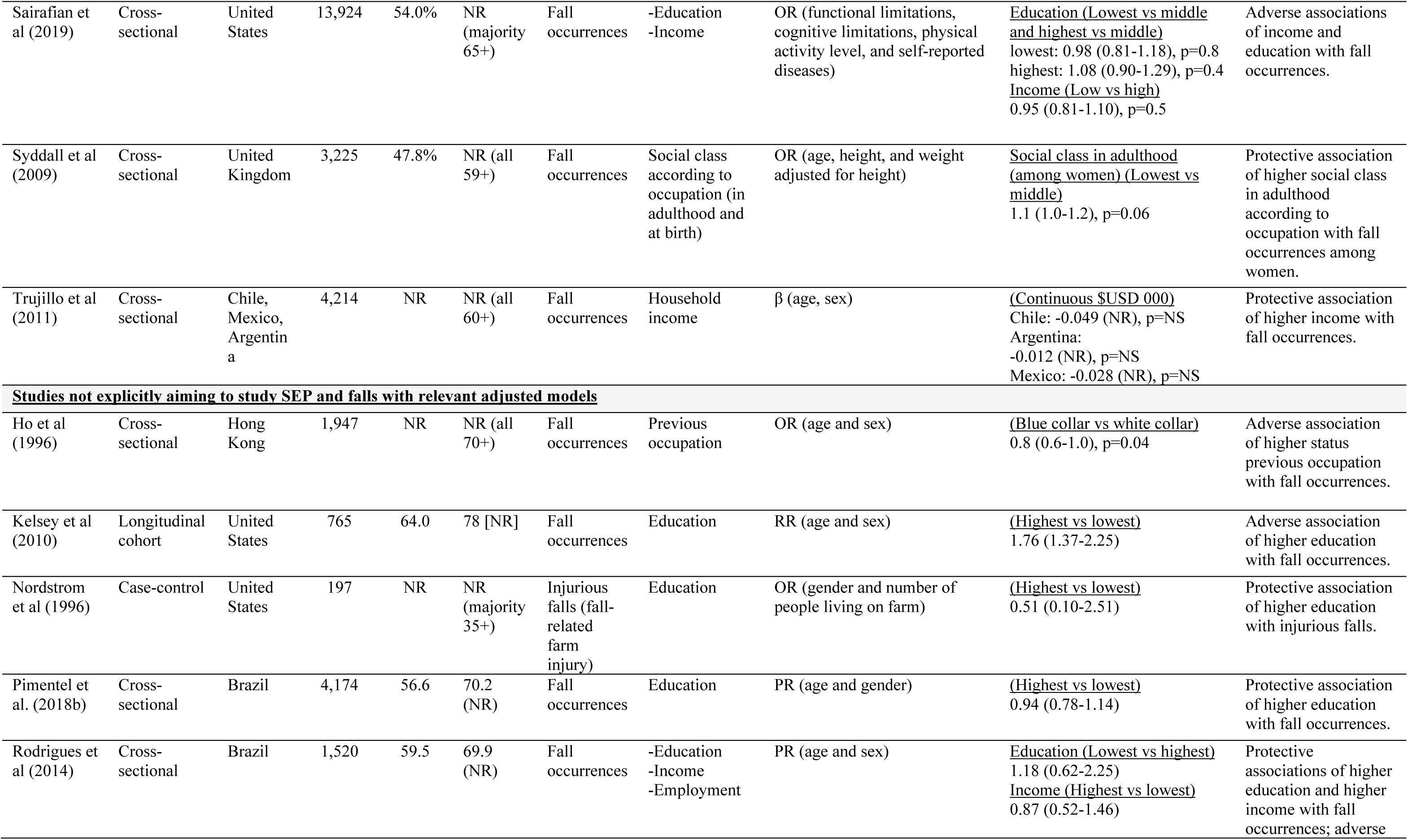
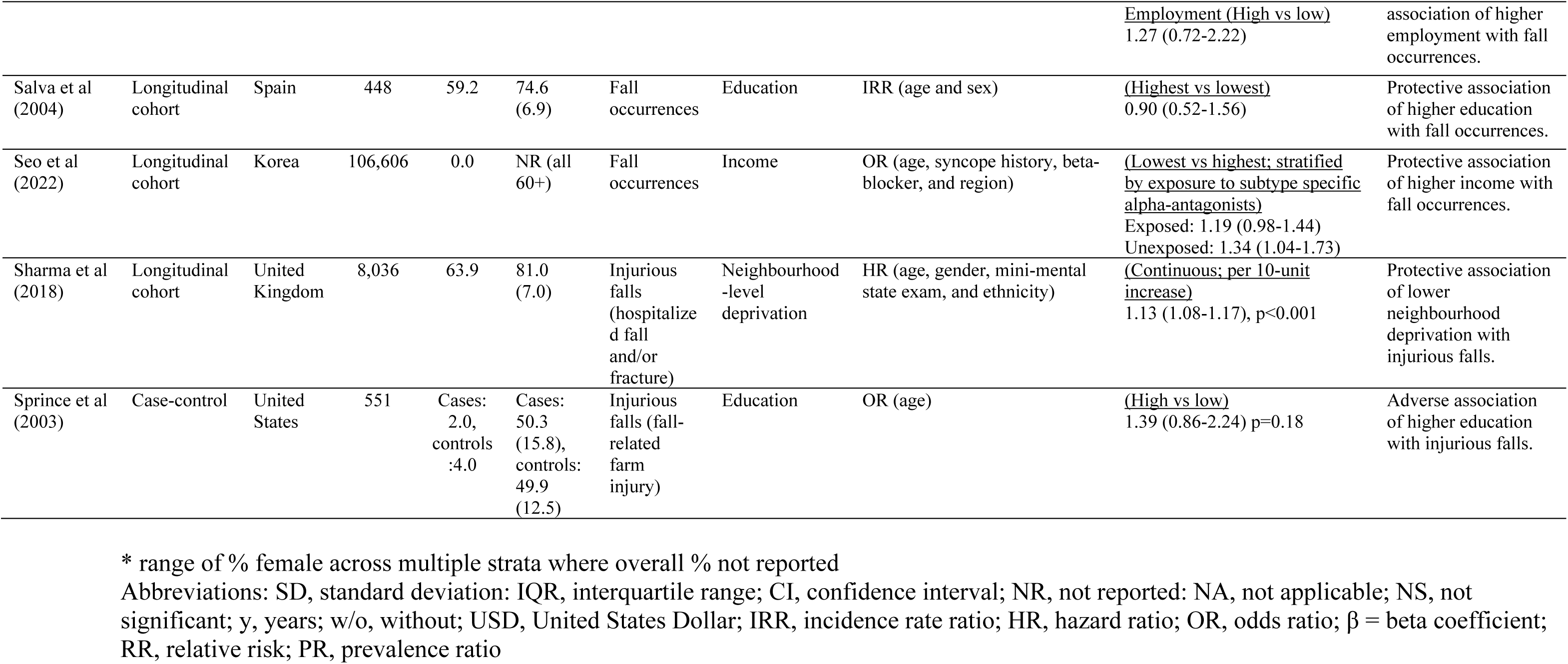
Characteristics of studies explicitly aiming to study SEP and falls and other studies with relevant adjusted models.

Fourteen studies explicitly aimed to study SEP and falls (Figure 2; Table 2) (27– 37,38(p202),39,40). Education was considered in five studies (three longitudinal cohort; two cross-sectional) (27,29,31,34,37). Protective associations between higher education and fall outcomes were reported in three studies (27,31,37), while the other two reported adverse associations.(29,34). Individual- and/or household-level income was analysed in five studies (two longitudinal cohort; three cross-sectional) (28,29,31,34,36). Higher income was protectively associated with fall outcomes in three studies (29,31,36) and adversely associated in one study (34), while the final study observed a protective association among middle-aged adults but an adverse association among older-aged adults (28). Occupation-related measures were examined in four studies (two longitudinal cohort; two cross-sectional) (29,31,35,37), where three found protective associations of higher-status occupations (29,35,37) and one found an adverse association (31). Only one longitudinal cohort study considered housing indicators as SEP measures, which reported a near-null adverse association between home ownership and fall occurrences (29). Individual- and/or household-level indexes of SEP measured based on education/income/occupation (29,31) or housing (33) were reported in three longitudinal cohort studies, all of which found protective associations between higher indexes of SEP and fall occurrences (29,31,33). Self-rated economic status was considered in one cross-sectional study that reported a protective association of this measure with fall occurrences (38), while another only reported unadjusted results on self-perceived economic situation (39). Neighbourhood-level SEP indicators were reported in three longitudinal cohort studies (30,32,40), which all generally reported protective associations between higher neighbourhood-level SEP and fall outcomes (30,32,40). Among all reported adverse associations between SEP indicators and falls, none were statistically significant while half of protective associations were statistically significant (Figure 2; Table 2).

**Figure 2.**
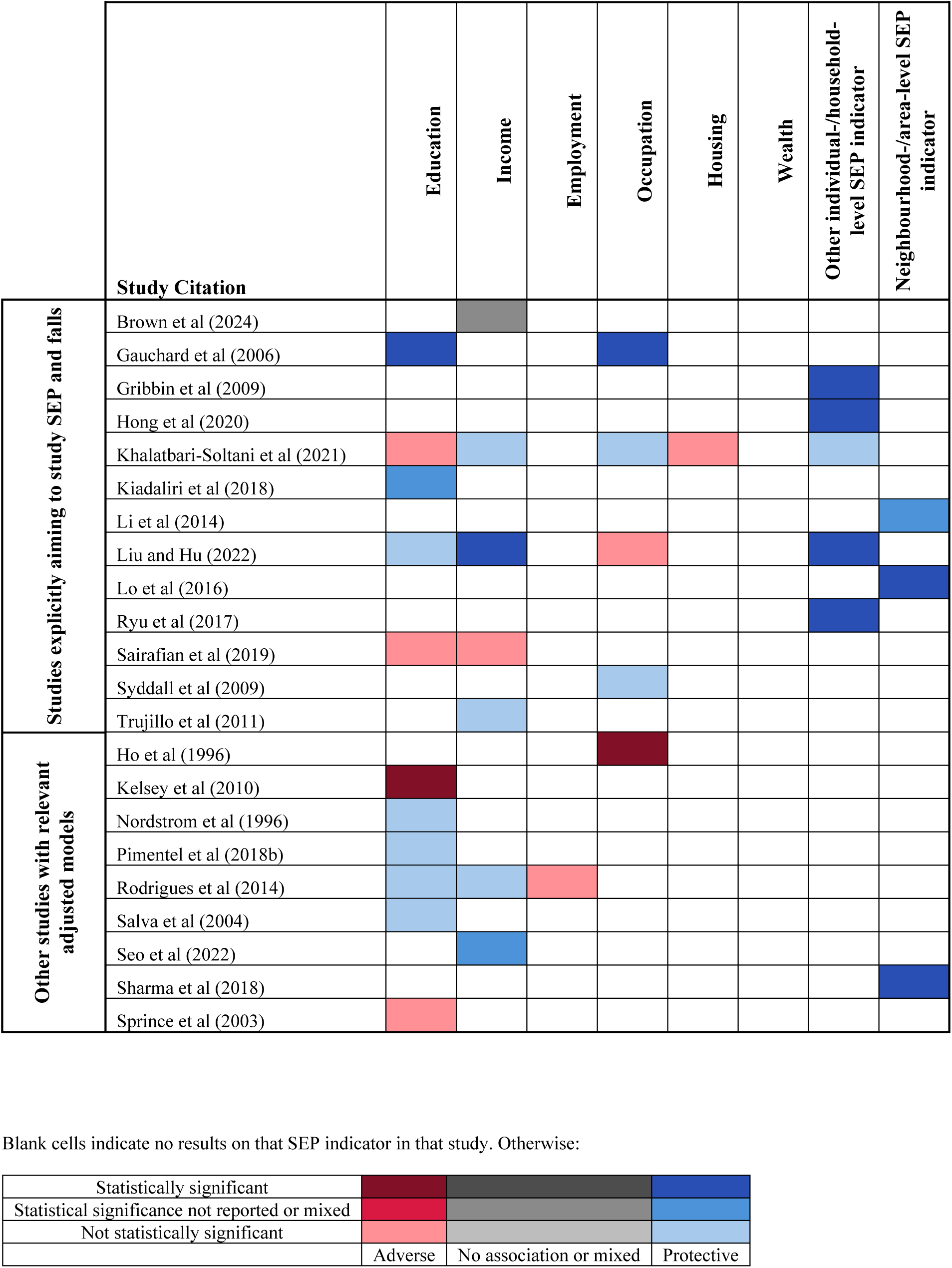
Visual overview of the associations between socioeconomic indicators and falls - results from relevant adjusted models.

Nine studies did not explicitly aim to study SEP and falls but reported relevant adjusted models (Figure 2; Table 2) (41–49). Education was analysed in six studies (42–45,48,49), of which four found a protective association between higher education and fall outcomes (43–45,49), and two reported an adverse association (42,48). Income was considered in two studies, both finding that higher income was protectively associated with fall occurrences (44,46). Occupation-related measures were reported in one study, where an adverse association of higher-status previous occupation with fall occurrences was found (41). Employment was considered in one study, which reported an adverse association between being employed and fall occurrences (44).

Neighbourhood-level SEP indicators were examined in one study, where lower deprivation was protectively associated with injurious falls (47). Of the adverse associations reported among these studies, half were statistically significant while only one protective association was statistically significant (Figure 2; Table 2).

In total, 102 studies did not explicitly aim to study SEP and falls and only contained relevant unadjusted results (Figure 3; Appendix pp16-36). Education was reported in 84 studies (50–133), with disproportionately high risks/rates of fall outcomes being found in low education groups compared to high education groups (low vs high for binary indicators and lowest vs highest for non-binary indicators) in 54 studies (50,52,54–58,61,64,66,67,69,70,72,73,76,78,81,83–85,85–88,92,96,98,100–102,104–108,110,112–116,118,120–124,126,127,129–131,133), and in high education groups compared to low education groups in 17 studies (59,62,63,74,75,77,79,80,82,97,99,103,109,111,117,125,128). Income was included in 27 studies (50,53,54,56,63,66,67,77,81,84,87,90,91,100,109,113,114,119,122,124,126,133–138), with falls being disproportionately experienced by low income groups in 18 studies (50,54,56,66,67,81,87,91,109,113,114,122,124,126,133,135,137,138) compared to five studies for the high income groups (63,77,84,134,136). Employment (e.g., employed vs unemployed, part-time vs full-time) was considered in 15 studies (52,64,69,80,94–96,98,100,104,108,116,123,125,137), ten of which found disproportionately high risks/rates of fall outcomes in the low employment group (52,64,69,95,96,98,104,108,123,137), while only three found higher risks/rates for the high employment group (80,100,116). Occupation was reported in six studies (51,79,87,88,132,139), where low status occupation groups faced a disproportionately high distribution of fall outcomes in three studies (87,88,139) compared to two for the high status occupation groups (51,132). For housing, all five studies with results on this SEP indicator found that lower housing groups (e.g., renter vs owner, poor household facilities) experienced disproportionately high rates/risks of fall outcomes (54,56,113,114,140). Wealth was included in three studies (61,112,116), with two finding disproportionately high risks/rates of falls in the low (112,116) and one study in the high (61) wealth groups. Other individual- and/or household-level SEP indicators were reported in 19 studies (57,64,69,83,102,108,113,114,118,121,123,141–148), 12 found fall outcomes to be disproportionately experienced by low SEP groups (57,83,113,114,118,123,141,142,144,146–148) compared to five studies for the high SEP groups (64,69,108,121,143). Neighbourhood- and/or area-level SEP was included in three studies (149–151), none of which found disproportionately high fall outcome risks/rates in the high SEP groups, while two found disproportionately high risks/rates in the low SEP groups (150,151). The remaining results were either mixed, unclear, or indicated no difference in the risk/rate of fall outcomes (51,53,60,65,68,71,79,90,93–95,100,102,119,125,132,145,149).

**Figure 3.**
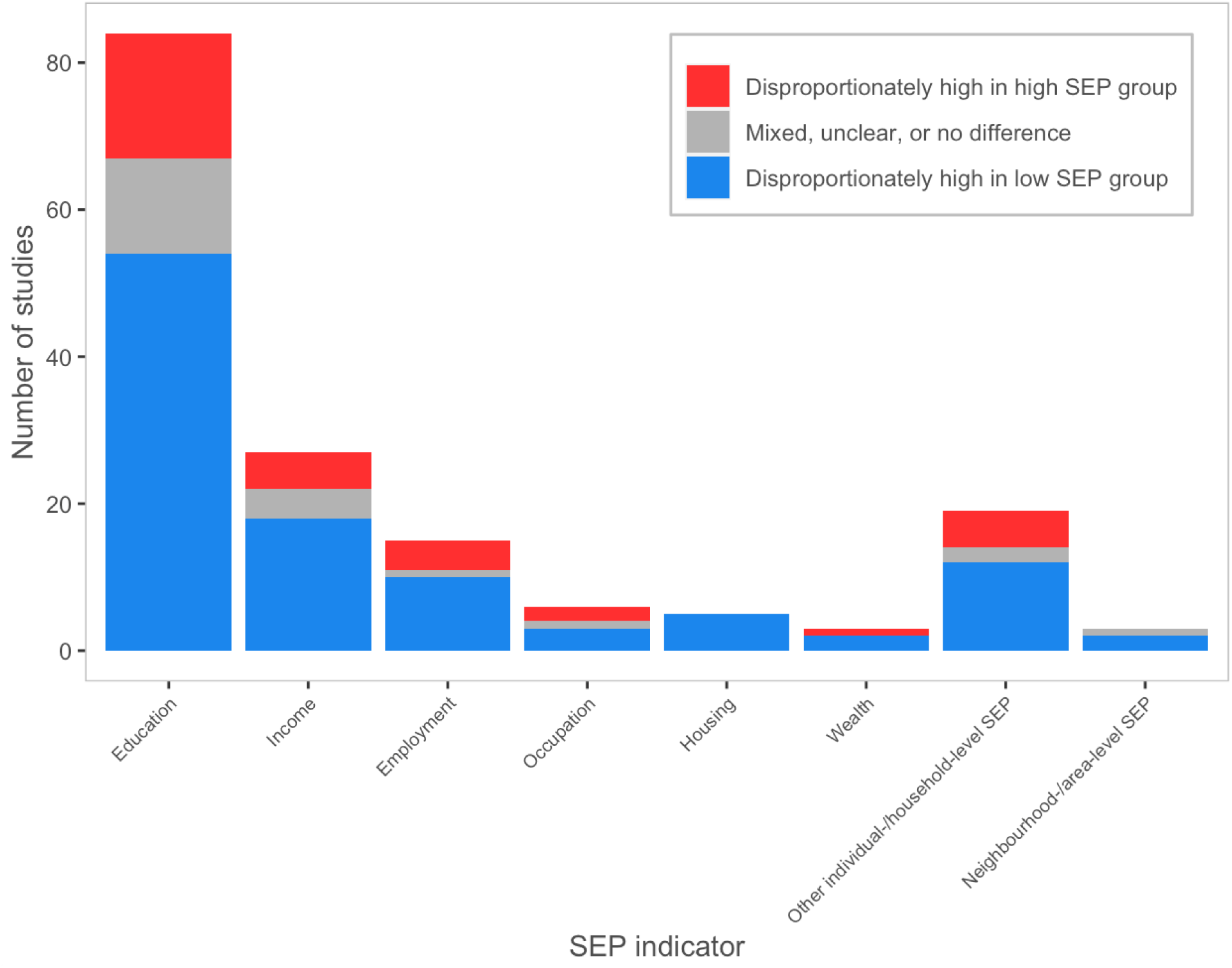
**Visual overview of results from studies not explicitly aiming to study socioeconomic position and falls with only relevant unadjusted results** Abbreviations: SEP, socioeconomic position. Created with R version 4.3.1

Only four of the 125 included studies had any results on SEP exposures from a stage of the life course prior to the time period of the study (29,31,35,41), of which three explicitly aimed to study SEP and falls (29,31,35). Three studies focused on associations of previous occupations with fall outcomes in adjusted models, as reported above (29,31,41). One of these models found a protective association between higher occupational position for the longest-held occupation in working life and fall occurrences at older ages (29). The other two models found an adverse association between higher-status past occupation and the occurrence of falls in older age (31,41). Only one study measured the same SEP indicator at multiple life course stages – social class measured based on father’s, husband’s, or individual’s occupational position (35).

Unadjusted results stratified by sex indicated that people of the lowest social class in adulthood experienced a disproportionately high risk/rate of fall occurrences in both sexes. However, for social class at birth, the distribution of fall occurrences was disproportionately high in the lowest social class group for men while being disproportionately high in the highest social class group for women (35). This suggests that social class exposures at varying life course periods may impact falls among women differently. However, the study did not explicitly test or discuss any life course conceptual models. Only one study explicitly evaluated the accumulation model and reported that higher cumulative SEP based on education, occupation, and income had a protective association with fall occurrences (29).

## Discussion

This review comprehensively synthesised existing literature on the relationship between SEP and falls among middle- and older-aged adults. Notably, while the number of included studies is substantial (n=125), only 14 of these explicitly aimed to study the relationship between SEP and falls, and only nine additional studies contained relevant adjusted models. A majority (60%-100%) of the estimates of effect from these 23 studies found a protective association between higher SEP and fall outcomes for various SEP indicators, except housing and employment where one non-statistically significant adverse association was found for each. Thus, the findings from these studies suggest that higher SEP generally has a protective relationship with falls. However, given the small number of such studies, the lack of statistical significance of many effect estimates, and several adverse associations having been observed, the overall body of evidence is limited. The findings from the 102 studies with results on the distribution of fall outcomes across SEP also show general socioeconomic inequities in fall outcomes, with 50% –100% of results for all SEP measures indicating that low SEP groups experience disproportionately high risks/rates of fall outcomes compared to high SEP groups.

The generally protective relationship between SEP and falls may reflect multifaceted mechanisms across a number of proposed pathways between SEP and health, including material, psychosocial, behavioural, biological, and health system factors (152). For example, one included study found that health shocks (i.e., illness and/or injury) explained a large part of the relationship between a SEP index and falls (31). Regarding behavioural factors, higher SEP is associated with increased leisure-time physical activity (153), and exercise is known to reduce the rate of falls (154). On the material pathway, higher SEP may provide access to better household environments (and resources to enable physical activity) – another known determinant of falls (155). Meanwhile, the observed adverse associations between higher SEP and falls may reflect study methods/designs, or a true adverse association between higher SEP and falls within specific study populations. For instance, in the case of occupation, an adverse association may be the result of people in higher status occupations engaging in lower rates of occupational physical activity (153), which may increase their risk of falls (154).

Notably, our results show that only four of the 125 included studies reported results that investigated the importance of timing and duration of being exposed to (dis)advantaged SEP and fall outcomes at older ages – all of which were occupation-related measures. Only one of these measured the same SEP indicator at two life stages – birth and adulthood. Unadjusted results from this study notably suggest a potentially differential impact of social class on fall occurrences at these two different life course stages among women (35). Further, only one of these studies provides results supporting the accumulation model, finding that higher cumulative SEP was protectively associated with fall occurrences (29). Overall, these results indicate a substantial knowledge gap regarding the impact of life course SEP on falls, which is concerning given the important knowledge that life course research provides to inform public health policies and interventions (10).

Future research is needed to address identified knowledge gaps on this topic. Studies need to cover a wide range of SEP measures to identify the specific populations in most need of targeted equity-focused fall prevention efforts, because individual socioeconomic measures have varied relationships to different health outcomes (156) with unique potential pathways through which they may impact health (6). Mediation analyses to understand the pathways between specific SEP variables and falls are also needed, as this can help identify intervention targets. Future research should also consider the role of timing, duration, and sequencing of exposures across the life course, which can inform the timing and targeting of interventions, including the life course periods where fall prevention efforts can be most effective (10). Additionally, studies should also consider the role of intersectionality by examining how social factors such as sex, gender and race/ethnicity may intersect with SEP to shape fall inequalities (157).

Despite the limited existing body of evidence, most studies report either a protective relationship between higher SEP and falls or that lower SEP groups generally experience disproportionately high risks/rates of fall-related outcomes. Public health efforts may therefore benefit from implementing equity-focused fall prevention interventions targeted toward individuals, households, and communities of lower SEP. Notably, existing evidence has clearly shown that physical activity-based interventions are particularly effective in reducing the incidence of falls among middle- and older-aged adults (154). Therefore, effective exercise programs such as balance and functional exercises and Tai Chi (154) should be promoted and made easily accessible to people of lower SEP. However, to be effective, program interventions must also tackle the material, logistical, and psychosocial barriers to physical activity that people with lower SEP often face (158).

Strengths of this review include a comprehensive search strategy, adoption of a life course perspective and use of robust model selection approaches informed by causal diagrams to minimise the risks of table 2 fallacy, confounding, and overadjustment biases (15,17,20). Further, we also appraised the risk of overadjustment bias in included studies, a common bias that is seldom considered in systematic reviews on socioeconomic health inequities (21).

There are some limitations, including that only 20% of titles and abstracts were independently screened by two reviewers. While we made efforts to select the most appropriate adjusted models of interest from included studies, a number of the results from these models are still subject to potential overadjustment bias (18,20,21). This emphasises the importance of causal diagrams being used in future research for appropriate aetiological model building (18). Our model and study selection approach also resulted in a majority of our results being in unadjusted form, which may be subject to confounding bias (26). Selective survival bias may also have impacted our results as inequitable premature death in younger ages among people of low SEP may lead to older low SEP populations comprising a relatively high proportion of healthy people, which can result in reductions and/or reversals of health inequalities (159,160). Finally, the QUIPS tool we used to appraise ROB may not effectively cover all important biases across different study types.

## Conclusion

Our review shows that only a limited number of studies have explicitly aimed to study the relationship between SEP and falls, and even fewer studies have studied the impact of life course SEP. While studies with relevant adjusted results generally found a protective association between higher SEP and falls, the overall body of evidence on this relationship is limited and subject to important biases. Studies only containing relevant unadjusted results also indicated that people of lower SEP experience a disproportionately high risk/rate of adverse fall outcomes compared to their high SEP counterparts. Equity-focused fall prevention interventions should therefore be considered, and future research is needed to inform interventions.

## Footnotes

### Author contributions

SKS conceptualised the study. FS, SKS, AvZ, and CS designed the study, with critical feedback on the study design provided by SC and FB. FS executed the search strategy. FS and MRF screened results, undertook data extraction, and appraised studies’ risk of bias. FS synthesised results and wrote the first draft with critical input from AvZ, SKS, and CS. All authors provided critical feedback, revisions of the manuscript, and approval for publication.

### Data availability statement

All data are available in the supplementary information.

## Data Availability

All data are available in the supplementary information.

