## Supplementary Material for "Socioeconomic Position and Falls Among Middle- and Older-Aged Adults: A Systematic Review with a Life Course Approach"

### Table S1 – PRISMA 2020 checklist

| **Section and Topic** | **Item #** | **Checklist item** | **Location where item is reported** |
| --- | --- | --- | --- |
| **TITLE** | | |  |
| Title | 1 | Identify the report as a systematic review. | Title |
| **ABSTRACT** | | |  |
| Abstract | 2 | See the PRISMA 2020 for Abstracts checklist. | ·· |
| **INTRODUCTION** | | |  |
| Rationale | 3 | Describe the rationale for the review in the context of existing knowledge. | Introduction |
| Objectives | 4 | Provide an explicit statement of the objective(s) or question(s) the review addresses. | Introduction |
| **METHODS** | | |  |
| Eligibility criteria | 5 | Specify the inclusion and exclusion criteria for the review and how studies were grouped for the syntheses. | Methods (Eligibility criteria) |
| Information sources | 6 | Specify all databases, registers, websites, organisations, reference lists and other sources searched or consulted to identify studies. Specify the date when each source was last searched or consulted. | Methods (Search strategy) |
| Search strategy | 7 | Present the full search strategies for all databases, registers and websites, including any filters and limits used. | Methods (Search strategy) and Table S2 |
| Selection process | 8 | Specify the methods used to decide whether a study met the inclusion criteria of the review, including how many reviewers screened each record and each report retrieved, whether they worked independently, and if applicable, details of automation tools used in the process. | Methods (Study selection) |
| Data collection process | 9 | Specify the methods used to collect data from reports, including how many reviewers collected data from each report, whether they worked independently, any processes for obtaining or confirming data from study investigators, and if applicable, details of automation tools used in the process. | Methods (Data extraction) |
| Data items | 10a | List and define all outcomes for which data were sought. Specify whether all results that were compatible with each outcome domain in each study were sought (e.g. for all measures, time points, analyses), and if not, the methods used to decide which results to collect. | Methods (Data extraction) |
|  | 10b | List and define all other variables for which data were sought (e.g. participant and intervention characteristics, funding sources). Describe any assumptions made about any missing or unclear information. | Methods (Data extraction) |
| Study risk of bias assessment | 11 | Specify the methods used to assess risk of bias in the included studies, including details of the tool(s) used, how many reviewers assessed each study and whether they worked independently, and if applicable, details of automation tools used in the process. | Methods (Risk of bias assessment) and Table S3 |
| Effect measures | 12 | Specify for each outcome the effect measure(s) (e.g. risk ratio, mean difference) used in the synthesis or presentation of results. | Tables 2, 3, S9 |
| Synthesis methods | 13a | Describe the processes used to decide which studies were eligible for each synthesis (e.g. tabulating the study intervention characteristics and comparing against the planned groups for each synthesis (item #5)). | Methods (Data synthesis) |
|  | 13b | Describe any methods required to prepare the data for presentation or synthesis, such as handling of missing summary statistics, or data conversions. | Methods (Data synthesis) |
|  | 13c | Describe any methods used to tabulate or visually display results of individual studies and syntheses. | Tables 2, 3, and S9, Figures 2 and 3 |
|  | 13d | Describe any methods used to synthesize results and provide a rationale for the choice(s). If meta-analysis was performed, describe the model(s), method(s) to identify the presence and extent of statistical heterogeneity, and software package(s) used. | Methods (Data synthesis) |
|  | 13e | Describe any methods used to explore possible causes of heterogeneity among study results (e.g. subgroup analysis, meta-regression). | Methods (Data synthesis) |
|  | 13f | Describe any sensitivity analyses conducted to assess robustness of the synthesized results. | Methods (Data synthesis) |
| Reporting bias assessment | 14 | Describe any methods used to assess risk of bias due to missing results in a synthesis (arising from reporting biases). | Methods (Risk of bias assessment) |
| Certainty assessment | 15 | Describe any methods used to assess certainty (or confidence) in the body of evidence for an outcome. | Methods (Risk of bias assessment) |
| **RESULTS** | | |  |
| Study selection | 16a | Describe the results of the search and selection process, from the number of records identified in the search to the number of studies included in the review, ideally using a flow diagram. | Results and Figure 1 |
|  | 16b | Cite studies that might appear to meet the inclusion criteria, but which were excluded, and explain why they were excluded. | Table S8 |
| Study characteristics | 17 | Cite each included study and present its characteristics. | Tables 2, 3, S9 |
| Risk of bias in studies | 18 | Present assessments of risk of bias for each included study. | Methods (Risk of bias assessment), Table S10, and Appendix pp39-40 |
| Results of individual studies | 19 | For all outcomes, present, for each study: (a) summary statistics for each group (where appropriate) and (b) an effect estimate and its precision (e.g. confidence/credible interval), ideally using structured tables or plots. | Table 2, 3, and S9 |
| Results of syntheses | 20a | For each synthesis, briefly summarise the characteristics and risk of bias among contributing studies. | Results, Table 1, Table S10, and Appendix pp39-40 |
|  | 20b | Present results of all statistical syntheses conducted. If meta-analysis was done, present for each the summary estimate and its precision (e.g. confidence/credible interval) and measures of statistical heterogeneity. If comparing groups, describe the direction of the effect. | Results, Tables 2, 3, and S9, and Figures 2 and 3 |
|  | 20c | Present results of all investigations of possible causes of heterogeneity among study results. | Tables S4, S5, S6, and S7 |
|  | 20d | Present results of all sensitivity analyses conducted to assess the robustness of the synthesized results. | N/A |
| Reporting biases | 21 | Present assessments of risk of bias due to missing results (arising from reporting biases) for each synthesis assessed. | N/A |
| Certainty of evidence | 22 | Present assessments of certainty (or confidence) in the body of evidence for each outcome assessed. | Figure 2 and Appendix pp39-40 |
| **DISCUSSION** | | |  |
| Discussion | 23a | Provide a general interpretation of the results in the context of other evidence. | Discussion |
|  | 23b | Discuss any limitations of the evidence included in the review. | Discussion |
|  | 23c | Discuss any limitations of the review processes used. | Discussion |
|  | 23d | Discuss implications of the results for practice, policy, and future research. | Discussion |
| **OTHER INFORMATION** | | |  |
| Registration and protocol | 24a | Provide registration information for the review, including register name and registration number, or state that the review was not registered. | Abstract and Introduction |
|  | 24b | Indicate where the review protocol can be accessed, or state that a protocol was not prepared. | Abstract and Introduction |
|  | 24c | Describe and explain any amendments to information provided at registration or in the protocol. | Methods |
| Support | 25 | Describe sources of financial or non-financial support for the review, and the role of the funders or sponsors in the review. | Source of funding and Abstract |
| Competing interests | 26 | Declare any competing interests of review authors. | Declaration of interests |
| Availability of data, code and other materials | 27 | Report which of the following are publicly available and where they can be found: template data collection forms; data extracted from included studies; data used for all analyses; analytic code; any other materials used in the review. | Data sharing |

### Table S2 – Search strategy

**1) Socioeconomic position**

exp Socioeconomic Factors/ OR socioeconomic*.tw OR socio?economic*.tw OR income.tw OR wealth.tw OR education*.tw OR occupation*.tw OR employment.tw OR hous*.tw OR social disparit*.tw OR social inequ*.tw OR social class.tw OR social status.tw OR social position.tw

**AND 2) Falls**

Accidental Falls/ OR fall*.tw OR faller*.tw

**AND 3) Middle-/older-aged adults**

Exp Aged/ OR Middle Aged/ OR senior*.tw OR elder*.tw OR old*.tw OR aged.tw OR ag?ing.tw OR middle-age* OR middle?age*.tw OR mid?life.tw OR midlife.tw

**AND 4) Observational studies**

MEDLINE and Embase (inception to March 15^th^, 2024):

Observational Study/ OR exp Cohort Studies/ OR exp Case-Control Studies/ OR Cross-Sectional Studies/

PyscInfo (inception to March 15^th^, 2024):

observational*.tw OR cohort*.tw OR case?control*.tw OR cross?sectional*.tw OR longitudinal*.tw OR prospective*.tw OR retrospective*.tw

### Table S3 – Modified Quality in Prognostic Studies (QUIPS) tool

| **Biases** | **Issues to consider for judging overall rating of "Risk of bias"** | **Study Methods & Comments** | **Rating of reporting** | **Rating of "Risk of bias"** |
| --- | --- | --- | --- | --- |
| Instructions to assess the risk of each potential bias: | These issues will guide your thinking and judgment about the overall risk of bias within each of the 6 domains. Some 'issues' may not be relevant to the specific study or the review research question. These issues are taken together to inform the overall judgment of potential bias for each of the 6 domains. | Provide comments or text exerpts in the white boxes below, as necessary, to facilitate the consensus process that will follow. | Determine adequacy of reporting (yes, partial, no or unsure) | Determine potential risk of bias for each domain (High, Moderate, or Low considering all relevant issues) |
| **1. Study Participation** | **Goal: To judge the risk of selection bias (likelihood that relationship between exposure and outcome is different for participants and eligible non-participants).** |  |  |  |
| Source of target population | The source population or population of interest is adequately described for key characteristics (LIST). |  |  |  |
| Method used to identify population | The sampling frame and recruitment are adequately described, including methods to identify the sample sufficient to limit potential bias (number and type used, e.g.., referral patterns in health care) |  |  |  |
| Recruitment period | Period of recruitment is adequately described |  |  |  |
| Place of recruitment | Place of recruitment (setting and geographic location) are adequately described |  |  |  |
| Inclusion and exclusion criteria | Inclusion and exclusion criteria are adequately described (e.g., including explicit diagnostic criteria or “zero time” description). |  |  |  |
| Adequate study participation | There is adequate participation in the study by eligible individuals |  |  |  |
| Baseline characteristics | The baseline study sample (i.e., individuals entering the study) is adequately described for key characteristics (LIST). |  |  |  |
| **Summary Study Participation** | **The study sample represents the population of interest on key characteristics, sufficient to limit potential bias of the observed relationship between exposure and outcome.** |  |  |  |
| **2. Study Attrition** | **Goal: To judge the risk of attrition bias (likelihood that relationship between exposure and outcome are different for completing and non-completing participants).** |  |  |  |
| Proportion of baseline sample available for analysis | Response rate (i.e., proportion of study sample completing the study and providing outcome data) is adequate. |  |  |  |
| Attempts to collect information on participants who dropped out | Attempts to collect information on participants who dropped out of the study are described. |  |  |  |
| Reasons and potential impact of subjects lost to follow-up | Reasons for loss to follow-up are provided. |  |  |  |
| Outcome and exposure variable information on those lost to follow-up | a) Participants lost to follow-up are adequately described for key characteristics (LIST). b) There are no important differences between key characteristics (LIST) and outcomes in participants who completed the study and those who did not. |  |  |  |
| **Study Attrition Summary** | **Loss to follow-up (from baseline sample to study population analyzed) is not associated with key characteristics (i.e., the study data adequately represent the sample) sufficient to limit potential bias to the observed relationship between exposure and outcome.** |  |  |  |
| **3. Exposure Measurement** | **Goal: To judge the risk of measurement bias related to how outcome was measured (differential measurement of outcome related to the level of outcome).** |  |  |  |
| Definition of the exposure | A clear definition or description of exposure is provided (e.g., including dose, level, duration of exposure, and clear specification of the method of measurement). |  |  |  |
| Valid and reliable measurement of exposure | a) Method of exposure measurement is adequately valid and reliable to limit misclassification bias (e.g., may include relevant outside sources of information on measurement properties, also characteristics, such as blind measurement and limited reliance on recall). b) Continuous variables are reported or appropriate cut-points (i.e., not data-dependent) are used. |  |  |  |
| Method and setting of exposure measurement | The method and setting of measurement of exposure is the same for all study participants. |  |  |  |
| Proportion of data on exposure available for analysis | Adequate proportion of the study sample has complete data for exposure variable. |  |  |  |
| Method used for missing data | Appropriate methods of imputation are used for missing exposure data. |  |  |  |
| **Exposure Measurement summary** | **Exposure is adequately measured in study participants to sufficiently limit potential bias.** |  |  |  |
| **4. Outcome Measurement** | **Goal: To judge the risk of bias related to the measurement of outcome (differential measurement of outcome related to the baseline level of exposure).** |  |  |  |
| Definition of the outcome | A clear definition of outcome is provided, including duration of follow-up and level and extent of the outcome construct. |  |  |  |
| Valid and reliable measurement of outcome | The method of outcome measurement used is adequately valid and reliable to limit misclassification bias (e.g., may include relevant outside sources of information on measurement properties, also characteristics, such as blind measurement and confirmation of outcome with valid and reliable test). |  |  |  |
| Method and setting of outcome measurement | The method and setting of outcome measurement is the same for all study participants. |  |  |  |
| **Outcome Measurement Summary** | **Outcome of interest is adequately measured in study aprticiapnts to sufficiently limit potential bias** |  |  |  |
| **5. Study Confounding** | **Goal: To judge the risk of bias due to confounding (i.e. the effect of exposure is distorted by another factor that is related to PF and outcome).** |  |  |  |
| Important confounders measured | All important confounders, including treatments (key variables in conceptual model: LIST) are measured. |  |  |  |
| Definition of the confounding factor | Clear definitions of the important confounders measured are provided (e.g., including dose, level, and duration of exposures). |  |  |  |
| Valid and reliable measured of confounders | Measurement of all important confounders is adequately valid and reliable (e.g., may include relevant outside sources of information on measurement properties, also characteristics, such as blind measurement and limited reliance on recall). |  |  |  |
| Method and setting of confounding measurement | The method and setting of confounding measurement are the same for all study participants. |  |  |  |
| Method used for missing data | Appropriate methods are used if imputation is used for missing confounder data |  |  |  |
| Appropriate accounting for confounding | a) Important potential confounders are accounted for in the study design (e.g., matching for key variables, stratification, or initial assembly of comparable groups). b) Important potential confounders are accounted for in the analysis (i.e., appropriate adjustment). |  |  |  |
| **Study confounding summary** | **Important potential confounders are appropriately accounted for, limiting potential bias with respect to the relationship between exposure and outcome.** |  |  |  |
| **6. Statistical analysis and reporting** | **Goal: To judge the risk of bias related to the statistical analysis and presentation of results.** |  |  |  |
| Presentation of analytical stratgy | There is sufficient presentation of data to assess the adequacy of the analysis. |  |  |  |
| Model development strategy | a) The strategy for model building (i.e., inclusion of variables in the statistical model) is appropriate and is based on a conceptual framework or model. b) The selected statistical model is adequate for the design of the study. |  |  |  |
| Reporting of results | There is no selective reporting of results |  |  |  |
| **Statistical analysis and presentation summary** | **The statistical analysis is appropriate for the design of the study, limiting potential for presentation of invalid or spurious results.** |  |  |  |
| **7. Overadjustment bias** | **Goal: To judge the risk of bias related to overadjustment (ie. introduction of bias through innapropriate model adjustments)** |  |  |  |
| Overadjustment factors | Did the authors control for any variables after the start of the exposure period being studies that could have been affected by the exposure? |  |  |  |
| Time-varying overadjustment factors | Did the authors control for time-varying factors or other varaibles measured after the start of the exposure window being studied? |  |  |  |
| **Overadjustment bias summary** | **Important mediating and collider variables are not inappropriately adjusted for, limiting the risk of overadjustment bias in the relationship between exposure and outcome.** |  |  |  |

### Table S4 – Measurement of socioeconomic indicators among studies explicitly aiming to study SEP and falls

| **Indicators of SEP** | **Number of studies** | **References** |
| --- | --- | --- |
| **Education measures**  1. Years of education   - Categorial   - > 3 levels   2. Education level   - Categorical   - 3 levels     - University, other post-qualification degree, no post-qualification degree     - University, middle and high-schools, primary school     - High school and above, junior high school graduation, primary school graduation or below     - More than high school, high school, less than high school | **5**  1  1  1  1  1 | Kiadaliri et al (2018)  Khalatbari–Soltani et al (2021)  Gauchard et al (2006)  Liu and Hu (2022)  Sairafian et al (2019) |
| **Income measures**  1. Household/family income   - Categorical   - 3 levels   - > 3 levels - Continuous   2. Individual income   - Categorical   - 2 levels   - 3 levels | **5**  1  1  1  1  1 | Liu and Hu (2022)  Brown et al (2024)  Trujillo et al (2011)  Sairafian et al (2019)  Khalatbari–Soltani et al (2021) |
| **Occupation measures**  1. Occupation   - Categorical   - 6 categories   - 7 categories   2. Previous occupation   - Categorical   - 3 levels     - High, intermediate, low     - Senior, middle, ordinary | **4**  1  1  1  1 | Syddall et al (2009)  Gauchard et al (2006)  Khalatbari–Soltani et al (2021)  Liu and Hu (2022) |
| **Housing measures**  1. Home ownership   - Categorical   - 2 levels | **1**  1 | Khalatbari–Soltani et al (2021) |
| **Other individual-/family-level indicators**  1. Indexes of socioeconomic position   - Categorical   - 3 levels   - > 3 levels - Continuous   2. Self-rated economic status   - Categorical   - > 3 levels | **4**  1  1  1  1 | Khalatbari–Soltani et al (2021)  Ryu et al (2017)  Liu and Hu (2022)  Hong et al (2020) |
| **Neighbourhood-level indicators**  1. Deprivation/disadvantage   - Categorical   - > 3 levels - 4 levels (quartiles)   2. Income   - Per-capita income ($1000s) - % per-capita income < poverty line   3. Housing   - % owner-occupied housing - % housing units vacant   4. Employment   - % adults who work   5. Social assistance   - % on public assistance | **3**  1  1  1  1  1  1  1  1 | Gribbin et al (2009)  Lo et al (2016)  Li et al (2014)  Li et al (2014)  Li et al (2014)  Li et al (2014)  Li et al (2014)  Li et al (2014) |

### Table S5 – Measurements of fall outcomes among studies explicitly aiming to study SEP and falls

| **Indicators of Falls** | **Number of studies** | **References** |
| --- | --- | --- |
| **Fall occurrences**  1. Self-reported   - Past year - Past 6 months - Past 4 months - Past 1 month - Timing unclear   2. Other ascertainment method | **9**  4  1  1  1  1  1 | Lo et al (2016), Sairafian et al (2019), Syddall et al (2009), Trujillo et al (2011)  Hong et al (2020)  Khalatbari–Soltani et al (2021)  Li et al (2014)  Liu and Hu (2022)  Gribbin et al (2009) |
| **Injurious falls**  1. Self-reported  2. Other ascertainment method | **2**  1  1 | Gauchard et al (2006)  Ryu et al (2017) |
| **Recurrent falls**  1. Self-reported  2. Other ascertainment method | **2**  1  1 | Brown et al (2024)  Gribbin et al (2009) |
| **Fall-related death** | **1** | Kiadaliri et al (2018) |

### Table S6 – Measurement of socioeconomic indicators among studies that did not explicitly aim to study SEP and falls but contained relevant adjusted results

| **Indicators of SEP** | **Number of studies** | **References** |
| --- | --- | --- |
| **Education measures**  1. Years of education   - Categorical   - 4 levels     - Never studied, 1-4, 5-8, 9+     - 0-3, 4-7, 8-11, 12+   - 5 levels - Continuous   2. Education level   - Categorical   - 2 levels   - 3 levels | **6**  1  1  1  1  1  1 | Pimentel et al (2018b)  Rodrigues et al (2014)  Nordstrom et al (1996)  Salva et al (2004)  Sprince et al (2003)  Kelsey et al (2010) |
| **Income measures**  1. Household/family income   - Categorical   - 4 levels   2. Individual income   - Quartiles | **2**  1  1 | Rodrigues et al (2014)  Seo et al (2022) |
| **Employment measures** | **1** | Rodrigues et al (2014) |
| **Occupation measures** | **1** | Ho et al (1996) |
| **Neighbourhood-level indicators** | **1** | Sharma et al (2018) |

### Table S7 – Measurements of fall outcomes among studies that did not explicitly aim to study SEP and falls but contained relevant adjusted results

| **Indicators of Falls** | **Number of studies** | **References** |
| --- | --- | --- |
| **Fall occurrences**  1. Self-reported   - Past year - Past 90 days - Past 1 month | **6**  4  1  1 | Ho et at (1996), Pimentel et al (2018b), Rodrigues et al (2014), Salva et al (2004)  Seo et al (2022)  Kelsey et al (2010) |
| **Injurious falls**  1. Hospitalized fall and/or fracture  2. Fall-related farm injury | **3**  1  2 | Sharma et al (2018)  Nordstrom et al (1996), Sprince et al (2003) |

### Figure S1 – Causal diagrams

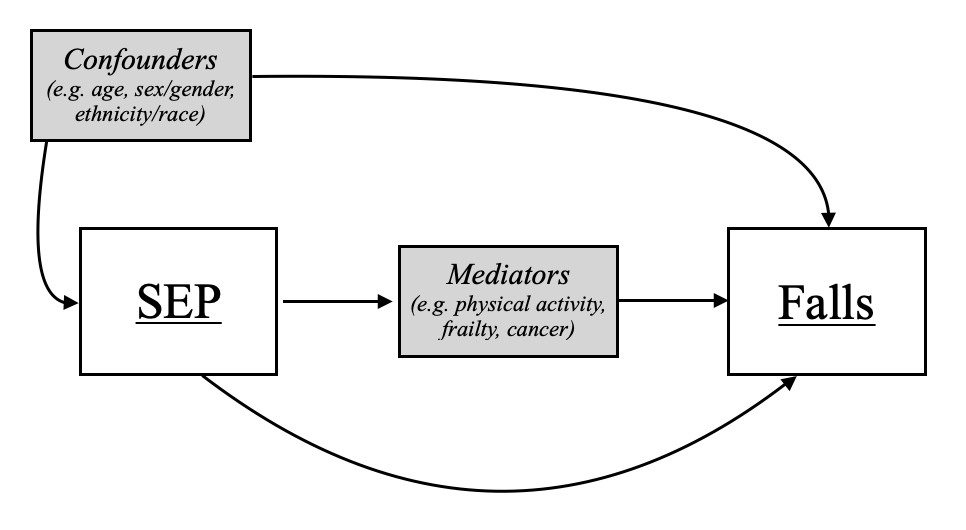

A) *Simplified causal diagram of the association between SEP and falls*

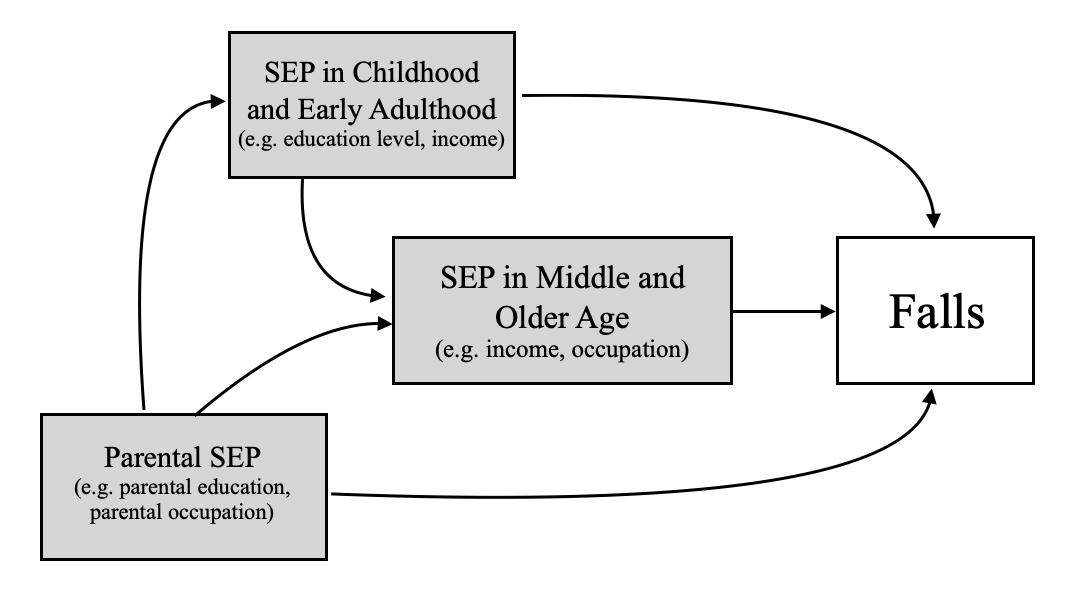

B) *Simplified causal diagram of the association between SEP at different periods of the life course and falls*

### Table S8 – Studies excluded during full-text screening with reason for exclusion

**Not a study design of interest (N = 35)**

Barrett-Connor et al., 2009; Casteel et al., 2018; Castro V.M. et al., 2014; Chen, He, et al., 2023; Chen, Lin, et al., 2023; Engchuan et al., 2019; Filshtein et al., 2019; Finlayson et al., 2006; Gokcek M.B. et al., 2019; Hayashi et al., 2023; Helgadottir et al., 2015; Hill et al., 2011, p. 20; Hiorth Y.H. et al., 2014; Hollinghurst J. et al., 2022; Jia H. et al., 2019; Jones et al., 2022; Lage I. et al., 2023; J. E. Lee & Stokic, 2008; Lima Rebelo et al., 2022; Maruf et al., 2016; Miro et al., 2018; Nugraha S. et al., 2022; Paiva E.P. et al., 2022; Perry et al., 2012; Pieper et al., 2012; Pirrie et al., 2020; Raina et al., 2016, p. 20; Ramirez-Martinez L. et al., 2020; Risbridger et al., 2022; Rondeau, 2022; Scheetz, 2015; van de Loo et al., 2022; Vincent-Onabajo G. et al., 2016; Zhang L. et al., 2022; Zou M. et al., 2023 (1-35)

**Not a publication type of interest (N = 30)**

Abbs E. et al., 2019; Ain S.N. et al., 2019; Aqlan M. et al., 2022; Blackwood J. et al., 2016; Catikkas N.M. et al., 2023; CHEN C.H. et al., 2019; Darling A. et al., 2018; Gade G.V. et al., 2022; Galica A. et al., 2010; Gardea Resendez M. et al., 2018; Hoffman & Rodriguez, 2015; Huang M.H. et al., 2016b, 2016a; Khalagi K. et al., 2022; Kim K.M. et al., 2023; Levett T. et al., 2016; Loo B.V.D. et al., 2020; Mamun et al., 2017; Midgley W. et al., 2023; Nguyen D.P. et al., 2021; Nume A. et al., 2017; Pang G.H.M. et al., 2019; Pastor Ochoa C. et al., 2011; Sanchez F.J.M. et al., 2018; Sharma A. et al., 2023; Shi J. et al., 2016; Stubbs B., 2019; Taseh A. et al., 2024; Verma V. et al., 2023; Welch S.A. et al., 2019 (36-65)

**Does not include the outcome of interest (N = 17)**

Akosile C.O. et al., 2023; Cross S.H. et al., 2022; Heybeli et al., 2022; Hwang et al., 2011; Johnson & Dias, 2019; Kempen et al., 2003; Lee S.M. et al., 2017; Y.-C. Lee et al., 2022; Moreland B.L. et al., 2018; Okuyama et al., 2020; Piazzalunga D. et al., 2020; Saengsuwan et al., 2014; Smith et al., 2017; Stel et al., 2003; Tan et al., 2016; Wei et al., 2010; Zille de Queiroz et al., 2020 (66-82)

**Does not include an indicator of the exposure of interest (N = 78)**

Abou L. & Rice L.A., 2022; Andresen et al., 2006; Bagi H.R.M. et al., 2019; Barbosa A.D.S. et al., 2019; Bekelis et al., 2015; Bhangu J. et al., 2017; Chihuri S. & Wong C.K., 2018; Chippendale et al., 2017; Cook A. et al., 2023; Coutinho et al., 2009; Coutinho E.S.F. et al., 2012; Cox D.D. et al., 2022; Cruz D.M.C.D. et al., 2017; Deprey, 2009; Divani et al., 2009; Escosura Alegre et al., 2023; Formiga F. et al., 2007; Francois C. et al., 2017; Gashaw et al., 2020; Gazibara et al., 2015; Girard et al., 2014; Gomes et al., 2013; Gray & Hildebrand, 2000, p. 200; Hakkenbrak N.A.G. et al., 2020; Halawa O. et al., 2021; Hamdard K. et al., 2023; Harrison B. et al., 2001; Hayakawa T. et al., 2014; Heikkila et al., 2022; Hendrich A. et al., 1995; Hreha K. et al., 2023; H.-C. Huang et al., 2003; J.-W. Huang et al., 2017; Jonsson A.-C. et al., 2021; Kamel et al., 2013; Kelsey et al., 2012; Kong et al., 2014; Lecky F.E. et al., 2021; Leclerc et al., 2008, 2009; Lukaszyk et al., 2018; Mackenzie et al., 2002; Maggi et al., 2018; Martuchi S.D. et al., 2020; Matsumoto et al., 2012; Matsumoto et al., 2014; Moreira N.B. et al., 2018; Najafpour et al., 2019; Neziraj et al., 2021; Ng et al., 2017; Ogliari et al., 2022; O’Loughlin et al., 1993; Olsson Moller U. et al., 2013; Ong et al., 2022; Paiva et al., 2022; Pereira et al., 2017; Phonthee et al., 2013; Porto J.M. et al., 2020; Prathapan S. et al., 2017; Qi et al., 2023; Sasidharan et al., 2020; Schmid A.A. et al., 2010; Scott et al., 2018; Silva et al., 2012; Sohng et al., 2004; Spoelstra et al., 2010; Tanaka et al., 2018; Tiensoli S.D. et al., 2019; Tripathy et al., 2015; Turnbull et al., 2022; Wasserstein D. et al., 2013; Wu et al., 2013; Yadollahi M. et al., 2019; Yeoh et al., 2013; Yip W.F. et al., 2023; Zevallos-Ventura A.S. et al., 2020; Zhang H. et al., 2022; Zhang L. et al., 2019 (83-160)

**Not the population of interest (N = 10)**

Berner K. et al., 2019; Brooks et al., 2023; Choi et al., 2014; Grivna et al., 2014; Lombardi et al., 2011; Mondal J. & Singh M., 2023; Mosenthal et al., 1995; Saadat et al., 2016; Sayyah et al., 2013; Stewart et al., 2016 (161-170)

**Full-text cannot be found (N = 4)**

Baranzini F. et al., 2008; Lopez J.C.C. & Palma O.A.V., 2023; Moskowitz G. et al., 2020; Yasumura et al., 1994 (171-174)

**Retracted article (N = 1)**

Soomar S.M. & Dhalla Z., 2023 (175)

### Table S9 – Characteristics of studies not explicitly aiming to study SEP and falls that contain relevant adjusted models

| Author, Publication date | Study type | Country | Sample size | % Female | Mean (SD) or Median [IQR] age | Fall outcome(s) | SEP Measures | Method of reporting | Results | Main findings |
| --- | --- | --- | --- | --- | --- | --- | --- | --- | --- | --- |
| Abreu et al (2016) | Longitudinal cohort | Brazil | 109 | 65·0 | NR (all 60+) | Recurrent falls | -Education -Income | Unadjusted relative risk (95% CI) | Education (Low vs high) 1·53 (0·95-2·49), p=NR Income (Low vs high) 1·44 (0·88-1·34), p=NR | Disproportionately high risk/rate of recurrent falls in low education and low income groups. |
| Aburub et al (2023) | Cross-sectional | Canada, Colombia, Brazil, and Albania | 1,995 | 45·9-55·0* | 69·5 (2·8), 69·2 (2·9) † | Fall occurrences | -Education -Occupation | Within each fall group, % in each exposure group. (analysis stratified to only those with cardiovascular disease) | Education lowest: 46·6% fallers, 38·0% non-fallers highest: 43·5% fallers, 16·4% non-fallers p=NR Occupation low: 61·1% fallers, 63·7% non-fallers high: 38·9% fallers, 36·3% non-fallers p=NR | Unclear results regarding the distribution of fall occurrences by education group. Disproportionately high risk/rate of fall occurrences in high status occupation group. |
| Agudelo-Botero (2018) | Cross-sectional | Mexico | 9,598 | 48·6-59·7* | NR (all 60+) | -Falls occurrences -Recurrent falls | -Education -Employment | Unadjusted odds ratios (95% CI) | Education (Low vs high) fall occurrences: 1·27 (1·06-1·52) recurrent falls: 1·68 (1·44-1·97) p<0·001 (overall p-value) Employment (Low vs high) fall occurrences: 1·21 (1·07-1·36) recurrent falls: 1·62 (1·46-1·79) p<0·001 (overall p-value) | Disproportionately high risk/rate of fall occurrences and recurrent falls in low education and low employment groups. |
| Ahmed et al (2021) | Longitudinal cohort | Canada, Colombia, Brazil, and Albania | 1979 | 52·4 | 69·1 (2·9) | -Fall occurrences -Recurrent falls -Injurious falls | -Education -Income | For education, mean (SD) education in each fall group. For income, within each fall group, % in each exposure group. | Education fall occurrences/recurrent falls: one fall: 10·59 (5·98) recurrent falls: 9·67 (6·19) no falls: 9·48 (5·65) p=0·008 (overall p-value) injurious falls: serious injury: 9·92 (4·81) moderate injury: 9·17 (5·94) no injury: 12·24 (6·19) p<0·001 (overall p-value) Income fall occurrences/recurrent falls: lowest: 39·18% one fall, 49·38% recurrent falls, 43·95% no falls highest: 25·08% one fall, 25·75% recurrent falls, 22·71% no falls p=0·04 (overall p-value) injurious falls: lowest: 54·90% serious injury, 49·70% moderate injury, 28·32% no injury highest: 19·61% serious injury, 18·60% moderate injury, 39·88% no injury p<0·001 (overall p-value) | Mixed results regarding the distribution of risk/rate of falls between different fall outcomes for education and income. |
| Alamri et al (2023) | Cross-sectional | Saudi Arabia | 403 | 67·2 | NR (all 60+) | Fall occurrences | -Education -Income -Housing | Within each exposure group, % with fall outcome. | Education lowest: 56·7% fallers highest: 42·9% fallers p=0·003 (overall p-value) Income lowest: 61·3% fallers highest: 46·7% fallers p=0·002 (overall p-value) Housing  low: 59·2% fallers high: 44·9% fallers p=0·029 (overall p-value) | Disproportionately high risk/rate of fall occurrences in low education, low income, and low housing groups. |
| Alex et al (2020) | Cross-sectional | Malaysia | 1,362 | 55-64·2* | NR (all 55+) | Fall occurrences | Education | % primary education in each fall group. | fallers: 30·6% non-fallers: 24·8% p=0·047 | Disproportionately high risk/rate of fall occurrences in low education group. |
| Almegbel et al (2018) | Cross-sectional | Saudi Arabia | 1,182 | 53·9 | 68·8 (9·03) | Fall occurrences | -Education -Income -Housing | Within each exposure group, % with fall outcome. | Education  lowest: 56·4% fallers highest: 38·5% fallers p=NR Income lowest: 54% fallers highest: 43·4% fallers p=NR Housing  low: 58·5% fallers high: 49% fallers p=NR | Disproportionately high risk/rate of fall occurrences in low education, low income, and low housing groups. |
| Almeida et al (2019) | Cross-sectional | Brazil | 211 | 62·1 | 73 (NR) | Fall occurrences | -Education -Index of property | Unadjusted prevalence ratio (95% CI, p-value) | Education (Low vs high) 1·29 (0·81-2·08), p=0·28 Index of property (Lowest vs highest) 1·69 (1·02-2·80), p=0·04 | Disproportionately high risk/rate of fall occurrences in low education and low index of property groups. |
| Arkkukangas et al (2021) | Cross-sectional | Sweden | 13,151 | 48·6 | NR (all 70+) | Injurious falls | Education | Within each exposure group, % non-fallers. | lowest: 84·0% non-fallers highest: 94·3% non-fallers p=NR | Disproportionately high risk/rate of injurious falls in low education group. |
| Arphorn et al (2022) | Cross-sectional | Thailand | 419 | 53·5 | NR (all 40+) | Occupational falls | Education | Unadjusted odds ratios (95% CI, p-value) | (High vs low) 1·13 (0·72-1·78), p=0·605 | Disproportionately high risk/rate of occupational falls in high education group. |
| Bally et al (2023) | Cross-sectional | Netherlands, Italy, Spain | 890 | 48·9-57·5* | 84·2 (6·8), 77·8 (6·3) † | Fall occurrences | Education | Within each fall group, what % are in each education group. (stratified by hospitalized and community-dwelling older adults) | Hospitalized older adults: low: 85·1% fallers, 81·6% non-fallers high: 14·9% fallers, 18·4% non-fallers p=0·640 (overall p-value) Community-dwelling older adults: low: 79·1% fallers, 80·5% non-fallers high: 20·9% fallers, 19·5% non-fallers p=0·666 (overall p-value) | Mixed results regarding the distribution of risk/rate of fall occurrences by education depending on strata. |
| Barik et al (2022) | Cross-sectional | India | 28,567 | 51·9 | 68·7 (7·3) | Fall occurrences | -Education -Wealth | Unadjusted odds ratios (95%CI) | Education (Low vs high) 1·15 (1·02-1·29), p=NR Wealth (Highest vs lowest) 1·17 (0·98-1·39), p=NR | Disproportionately high risk/rate of fall occurrences in low education group. Disproportionately high risk/rate of fall occurrences in high wealth group. |
| Biderman et al (2002) | Longitudinal cohort | Israel | 283 | 58·0 | 71 (NR) | Fall occurrences | Education | Unadjusted relative risk (95% CI) | (High vs low) 1·93 (0·99-3·77), p=NS | Disproportionately high risk/rate of fall occurrences in high education group. |
| Brito et al (2014) | Cross-sectional | Brazil | 316 | 54·5 | 74·2 (NR) | Fall occurrences | -Education -Income | Unadjusted prevalence ratios (95% CI, p-value) | Education (Low vs high) 0·93 (0·63-1·38), p=0·748 Income (Lowest vs highest) 0·90 (0·47-1·73), p=0·295 | Disproportionately high risk/rate of fall occurrences in high education and high income groups. |
| Cevizci et al (2015) | Cross-sectional | Turkey | 1,001 | 55·0 | 74·1 (NR) | Fall occurrences | -Education -Employment -Social assurance | Within each exposure group, % with fall outcome. | Education low: 27·1% fallers high: 19·6% fallers p=0·020 (overall p-value) Employment low: 25·7% fallers high: 13·8% fallers p=0·194 (overall p-value) Social assurance low: 24·6% fallers high: 40·8% fallers p=0·011 (overall p-value) | Disproportionately high risk/rate of fall occurrences in low education and low employment groups. Disproportionately high risk/rate of fall occurrences in high social assurance group. |
| Coutinho et al (2008) | Case-control | Brazil | 500 | 78·0 | 75·5 (8·2), 75·3 (7·7) † | -Injurious falls (severe fracture following a fall) | Education | Within each fall group, % within each exposure category. | lowest: 41·6% fallers, 44·0% non-fallers highest: 2·4% fallers, 4·4% non-fallers p=NR | Unclear results regarding the distribution of injurious falls by education group. |
| Dai et al (2018) | Cross-sectional | Singapore | 10,009 | 48·6-63·4* | 58·9 (10·4) | -Fall occurrences -Recurrent falls | -Education -Income | Within each fall group, % in each exposure group. | Education lowest 32·1% fallers, 34·8% recurrent fallers, 21·8% non-fallers highest: 34·2% fallers, 29·0% recurrent fallers, 40·3% non-fallers p<0·001 (overall p-value) Income lowest: 84·4% fallers, 89·9% recurrent fallers, 76·5% non-fallers p<0·001 (overall p-value) | Disproportionately high risk/rate of fall occurrences and recurrent falls in low education and low income groups. |
| de Souza Moreira et al (2022) | Cross-sectional | Brazil | 834 | 56·4 | 69·3 (7·7) | -Fall occurrences | Socioeconomic status | Unadjusted odds ratio (95% CI, p-value) | (Low vs high) 3·51 (1·27-9·67), p=0·015 | Disproportionately high risk/rate of fall occurrences in low socioeconomic status group. |
| do Nascimento et al (2017) | Cross-sectional | Brazil | 1,188 | 60·1 | NR (all· 60+) | -Fall occurrences | -Education -Income | Within each exposure group, % non-fallers. | Education lowest: 63·29% non-fallers highest: 74·14% non-fallers p=0·2415 Income low: 68·82% non- fallers high: 72·46% non-fallers p=0·1973 | Disproportionately high risk/rate of fall occurrences in low education and low income groups. |
| dos Santos et al (2015) | Longitudinal cohort | Brazil | 280 | 68·2 | 71·6 (6·7) | -Fall occurrences -Recurrent falls | Education | Within each fall group, % in each exposure category | Fall occurrences: low: 44% fallers, 47% non-fallers high: 56% fallers, 53% non-fallers p=0·62 Recurrent falls:  low: 50% recurrent fallers, 43·6% non-recurrent fallers high: 50% recurrent fallers, 56·4% non-recurrent fallers p=0·33 (overall p-value) | Mixed results regarding the distribution of risk/rate of falls between different fall outcomes for education. |
| Drewes et al (2021) | Cross-sectional | Germany | 897 | 13·0 | 57 (6·7) | Fall occurrences | Socioeconomic status | Unadjusted odds ratio (95%CI) | (Continuous) 0·93 (0·89-0·98, NR) | Disproportionately high risk/rate of fall occurrences in low socioeconomic status group. |
| El Sayed et al (2023) | Cross-sectional | Egypt | 289 | NR | 60·6 (8·0), 67·6 (10·6) † | Fall occurrences | -Education -Employment -Family financial ability | Within each fall group, % in each exposure group. | Education lowest: 25·0% fallers, 14·0% non-fallers highest: 34·4% fallers, 47·9% non-fallers p=0·181 (overall p-value) Employment  lowest: 53·1% fallers, 34·6% non-fallers highest: 18·8% fallers, 43·2% non-fallers p=0·026 (overall p-value) Family financial ability lowest: 9·4% fallers, 10·9% non-fallers highest: 12·5% fallers, 10·1% non-fallers p=0·66 (overall p-value) | Disproportionately high risk/rate of fall occurrences in low education and low employment groups. Disproportionately high risk/rate of fall occurrences in high family financial ability group. |
| Eshkoor et al (2014) | Cross-sectional | Malaysia | 1,210 | NR | NR (all 60+) | Fall occurrences | Education | Prevalence of falls in each exposure category. | low: 18·5% fallers high: 14·7% fallers p=0·054 | Disproportionately high risk/rate of fall occurrences in low education group. |
| Ferreira et al (2022) | Cross-sectional | Portugal | 69 | 65·2 | 83·1 (8·9) | Fall occurrences | Education | Within each fall group, % in each exposure category | lowest: 53·3% fallers, 63·0% non-fallers highest: 0% fallers, 3·7% non-fallers p=0·373 (overall p-value) | Unclear results regarding the distribution of fall occurrences by education group. |
| Fikadu et al (2021) | Cross-sectional | Ethiopia | 331 | 97·1 | 40·9 (19·6) | Fall deaths | Occupation | Unadjusted odds ratio (95% CI) p-value | (Lowest vs highest) 2·92 (0·91-9·38) p=0·032 (overall p-value) | Disproportionately high risk/rate of fall deaths in low occupation group. |
| Frankenthal et al (2021) | Cross-sectional | Israel | 3,159 | 57·3 | 75·1 (6·2) | Fall occurrences | Education | Within each fall group, % in each exposure category | low: 62·9% fallers, 55·1% non-fallers high: 37·1% fallers, 44·9% non-fallers p<0·001 (overall p-value) | Disproportionately high risk/rate of fall occurrences in low education group. |
| Gade et al (2021) | Longitudinal cohort | Denmark | 241 | 66·4 | 82 [80-86] | Fall occurrences | Education | Unadjusted incidence rate ratio (95%CI, p-value) | (Highest vs lowest) 0·84 (0·25-2·03), p=0·73 | Disproportionately high risk/rate of fall occurrences in low education group. |
| Garcia-Rudolph et al (2023) | Longitudinal cohort | Spain | 2,341 | 34·6 | 50·6 [35·8-64·2] | Inpatient falls | Education | Unadjusted hazard ratio (95%CI, p-value) | (High vs low) 1·03 (0·72-1·49), p=0·841 | Disproportionately high risk/rate of inpatient falls in high education group. |
| Gashaw and Admass (2021) | Cross-sectional | Ethiopia | 320 | 41·6 | 55·9 (14·2) | Injurious falls | Income | Within each exposure group, % with fall outcome. | lowest: 25·5% injurious falls highest: 26·3% injurious falls | Disproportionately high risk/rate of injurious falls in high income group. |
| Goncalves et al (2008) | Cross-sectional | Brazil | 180 | 75·0 | 80·2 (NR) | Fall occurrences | Education | Relative risk (95% CI, p-value) | (High vs low) 1·01 (0·64-1·62), p=0·94 | Disproportionately high risk/rate of fall occurrences in high education group. |
| Grimm et al (2022) | Longitudinal cohort | United States | 3,773 | 55·1 | NR (majority 60+) | Fall occurrences | Income | Within each fall group, % in each exposure group. | lowest: 22·53% fallers, 19·1% non-fallers highest: 17·79% fallers, 22·16% non-fallers p=0·519 (overall p-value) | Disproportionately high risk/rate of fall occurrences in low income group. |
| Hartley et al (2023) | Longitudinal cohort | Ireland | 8,154 | 54·2 | 63·8 (9·8) | Recurrent falls | Education | Within each fall group, % in each exposure category | lowest: 36·2% recurrent fallers, 30·2% non-recurrent fallers (p=0·003) highest: 25·9% recurrent fallers, 29·7% non-recurrent fallers (p=0·062) | Disproportionately high risk/rate of recurrent falls in low education group. |
| Huang et al (2019) | Longitudinal cohort | United States | 1,097 | NR | 72·5 (6·3), 71·9 (5·8) † | Fall occurrences | -Education -Income | Unadjusted odds ratios (95%CI, p-value) (stratified by breast cancer and prostate cancer survivors) | Education (Highest vs lowest) breast cancer survivors: 1·29 (0·63-2·62), p=0·49 prostate cancer survivors: 1·69 (0·96-2·96), p=0·07 Income (Highest vs lowest) breast cancer survivors: 1·41 (0·73-2·72), p=NR prostate cancer survivors: 1·23 (0·71-2·13), p=0·46 | Disproportionately high risk/rate of fall occurrences in high education and high groups. |
| Hu et al (2015) | Longitudinal cohort | China | 3,092 | 100·0 | 75·0 (9·4) | Fall occurrences | Education | Within each exposure group, % with fall outcome. | lowest: 10·1% fallers highest: 6·0% fallers p<0·01 (overall p-value) | Disproportionately high risk/rate of fall occurrences in low education group. |
| Janakiraman et al (2019) | Cross-sectional | Ethiopia | 599 | 54·6 | 61·0 (20·0) | Fall occurrences | -Education -Occupation | For education, unadjusted odds ratio (95% CI) p-value. For occupation, within each fall group, % in each exposure group. | Education (Highest vs lowest) 1·58 (1·89-2·78), p=NR Occupation lowest: 19·4% fallers, 20·7% non-fallers highest: 5·9% fallers, 15·2% non-fallers p=NR | Disproportionately high risk/rate of fall occurrences in high education group. Unclear results regarding the distribution of risk/rate of fall occurrences by occupation group. |
| Jorgensen et al (2016) | Cross-sectional | Norway, Sweden | 224 | 23·0 | 49·6 (14·9) | Recurrent falls | -Education -Employment | Within each fall group, % in each exposure category | Education  lowest: 29% recurrent fallers, 36% non-recurrent fallers highest: 42% recurrent fallers, 32% non-recurrent fallers p=0·234 (overall p-value) Employment  lowest: 47% recurrent fallers, 63% non-recurrent fallers highest: 45% recurrent fallers, 33% non-recurrent fallers p=0·05 (overall p-value) | Disproportionately high risk/rate of recurrent falls in high education and high employment groups. |
| Kalula et al (2016) | Cross-sectional | South Africa | 837 | 76·5 | 74 (6·1), 75 (7·1), 74 (6·2) † | Fall occurrences | Household socioeconomic status | Mean (IQR) SES index in each fall group. | fallers: 8 (7-8) non-fallers 7 (6-8) p < 0·001 | Disproportionately high risk/rate of fall occurrences in high socioeconomic status group. |
| Kim et al (2022) | Longitudinal cohort | Korea | 1,414 | 49·2 | NR (majority 65+) | -Fall occurrences -Injurious falls (falls requiring medical treatment) | -Education -Income | Within each exposure group, % with fall outcome. | Education  Fall occurrences: lowest (8·9%), highest (2·8%) p=0·001 (overall p-value) Injurious falls: lowest (6·1%), highest (1·9%) p=0·017 (overall p-value) Income Fall occurrences: lowest (8·4%), highest (4·3%) p=0·271 (overall p-value) Injurious falls: lowest (6·5%), highest (2·4%) p=0·136 (overall p-value) | Disproportionately high risk/rate of fall occurrences and injurious falls in low education and low income groups. |
| Kuhirunyaratn et al (2013) | Case-control | Thailand | 333 | 60·4-68·5* | 69 [11], 68 [10] | Fall occurrences | Education | Within each fall group, what % are in each exposure group. | lowest: 7·2% fallers, 9·9% non-fallers highest: 5·4% fallers, 5·4% non-fallers p=0·839 (overall p-value) | Disproportionately high risk/rate of fall occurrences in high education group. |
| Kummari et al (2024) | Cross-sectional | India | 1,028 | 50·1 | NR (all· 60+) | Fall occurrences | -Education -Socioeconomic status | Within each exposure group, % with fall outcome. | Education low: 32·33% high: 21·51% p<0·001 (overall p-value) Socioeconomic status low: 29·32% high: 7·78% p<0·001 (overall p-value) | Disproportionately high risk/rate of fall occurrences in low education and low socioeconomic status groups. |
| Lamba et al (2022) | Longitudinal cohort | United States | 42,648 | 51·0-58·0* | 76·0 (6·0) | Fall occurrences | -Neighbourhood-level education -Neighbourhood-level income | Unadjusted hazard ratios (95%CI, p-value) | Neighbourhood-level education (Continuous) 1 (1·00-1·01), p<0·001 Neighbourhood-level income (Continuous) 1·02 (1·01-1·03), p<0·001 | No difference in the risk/rate of fall occurrences by neighbourhood-level education group. Disproportionately high risk/rate of fall occurrences in high neighbourhood-level income group. |
| Lavedan Santamaria et al (2015) | Cross-sectional | Spain | 640 | 60·3 | 81·5 (5·0) | Fall occurrences | -Education -Income | Within each fall group, what % are in each exposure group. | Education low: 37·6% fallers, 36·4% non-fallers high: 62·4% fallers, 63·6% non-fallers p=NS (overall p-value) Income low: 52·7% fallers, 56·2% non-fallers high: 47·3% fallers, 43·8% non-fallers p=NS (overall p-value) | Disproportionately high risk/rate of fall occurrences in low education and low income groups. |
| Lee et al (2021) | Longitudinal cohort | Taiwan | 232 | 74·0 | 70·5 (9·2) | Fall occurrences | Education | Within each fall group, what % are in each exposure group. | lowest: 62·5% fallers, 24·4% non-fallers highest: 14·1% fallers, 22·0% non-fallers p<0·001 (overall p-value) | Disproportionately high risk/rate of fall occurrences in low education group. |
| Lee et al (2020) | Case-control | Korea | 4,114 | 34·3-47·1* | 61 (15·0), 621 (13·9), 59·73 (14·8), 55·0 (13·5) † | Fall occurrences | Education | Within each fall group, what % are in each exposure group. (stratified by fall risk group) | High-risk group: lowest: 24·2% fallers, 22·9% non-fallers highest: 19·9% fallers, 30·0% non-fallers p=0·001 (overall p-value) Low-risk group: lowest: 20·7% fallers, 14·8% non-fallers highest: 28·9% fallers, 31·8% non-fallers p=0·331 (overall p-value) | Disproportionately high risk/rate of fall occurrences in low education group. |
| Lin et al (2023) | Cross-sectional | China | 1,629 | 62·3 | NR (all 65+) | -Fall occurrences -Injurious falls | -Education -Income -Occupation | Prevalence of fall outcomes in each exposure group. | Education  Fall occurrences:  lowest: 16·50% (95%CI: 8·31-30·09) highest: 14·36% (95%CI: 10·31-19·66) p<0·001 (overall p-value)  Injurious falls: lowest: 11·46% (95%CI: 5·50-22·35) highest: 7·92% (95%CI: 5·61-11·95) p<0·001 (overall p-value)  Income Fall occurrences:  lowest: 13·17% (95%CI: 9·52-17·94) highest: 11·46% (95%CI: 7·67-16·78) p<0·001 (overall p-value)  Injurious falls: lowest: 7·01% (95%CI: 4·31-11·21) highest: 6·39% (95%CI: 3·79-10·59) p<0·001 (overall p-value)  Occupation Fall occurrences:  lowest: 22·97% (95%CI: 19·59-26·73) highest: 14·79% (95%CI: 12·29-17·7) p<0·001 (overall p-value) Injurious falls: lowest: 13·23% (95%CI: 8·71-19·59) highest: 9·92% (95%CI: 7·71-12·69) p<0·001 (overall p-value) | Disproportionately high risk/rate of fall occurrences and injurious falls in low education, low income, and low occupation groups. |
| Lin et al (2022) | Cross-sectional | China | 5,374 | 55·7 | 69·3 (6·8) | Fall occurrences | -Education -Occupation | Within each exposure group, % with fall outcome. | Education  lowest: 14·8% highest: 10·7% p<0·001 (overall p-value) Occupation low: 12·9% high: 10·0% p=0·002 (overall p-value) | Disproportionately high risk/rate of fall occurrences in low education and low occupation groups. |
| Li et al (2021) | Cross-sectional | China | 16,240 | 53·1 | NR (majority 45-65) | Fall occurrences | Education | Prevalence of fall outcome in each exposure group. | lowest: 19·1% highest: 11·0% p<0·001 (overall p-value) | Disproportionately high risk/rate of fall occurrences in low education group. |
| Luukinen et al (1996) | Case-control | Finland | 1,016 | 61·0 | NR (all older adults) | Recurrent falls | Housing | Proportion in each fall category that have poor household facilities. | recurrent fallers: 35% controls: 28% p=NR | Disproportionately high risk/rate of recurrent falls in low housing group. |
| Maneeprom et al (2018) | Cross-sectional | Thailand | 64 | 81·2 | 76·4 (9·6) | Fall occurrences | -Education -Income | Proportion of entire study sample. | Education low: 29·7% fallers, 10·9% non-fallers high: 34·4% fallers, 25·0% non-fallers p=0·214 (overall p-value) Income low: 10·9% fallers, 3·1% non-fallers high: 53·1% fallers, 32·8% non-fallers p=0·355 (overall p-value) | Unclear results regarding the distribution of fall occurrences by education and income groups |
| Matsuda et al (2011) | Cross-sectional | United States | 474 | 82·7 | 51·1 (10·9) | -Fall occurrences -Injurious falls | -Education -Income | Unadjusted odds ratios (95%CI, p-value) | Education (Low vs high) fall occurrences: 0·81 (0·48-1·36), p=NS injurious falls: 1·04 (0·18-1·60), p=NS) Income (Low vs high) fall occurrences: 2·29 (1·44-3·67), p<0·05 injurious falls: 1·49 (0·75-2·96), p=NS | Mixed results regarding the distribution of risk/rate of falls between different fall outcomes for education. Disproportionately high risk/rate of injurious falls in low income group. |
| Menendez et al (2017) | Longitudinal cohort | United States | 328,874 | 58·8-60·9* | NR (majority 65+) | Inpatient falls | Neighbourhood-level median household income | Rate of fall outcome in each exposure category. | lowest: 15·3% highest: 11·2% p=NR | Disproportionately high risk/rate of inpatient falls in low neighbourhood-level income group. |
| Merchant et al (2023) | Cross-sectional | Singapore | 328 | 56·4 | 72·5 (5·7) | Fall occurrences | Education | Mean years of education (SD) in each fall group. | fall group: 6·3 years (4·2) non-fall group: 7·5 years (4·11) p=0·042 | Disproportionately high risk/rate of fall occurrences in low education group. |
| Mgabhi et al (2024) | Cross-sectional | Philippines, Vietnam | 8,984 | 55·3-59·6* | 67·8 (6·5), 69·53 (7·6) † | Fall occurrences | Education | Unadjusted odds ratios (95%CI, p-value). Stratified by country. | (Highest vs lowest) Philippines: 2·96 (2·37-3·68), p<0·001 Vietnam: 0·32 (0·10-1·02), p=0·054 | Mixed results regarding the distribution of falls between different countries by education group. |
| Moon et al (2021) | Longitudinal cohort | Korea | 8,263 | 48·8 | 73·84 (6·33) | Inpatient falls | Income | Unadjusted odds ratio (95%CI, p-value) | (Low vs high) 0·779 (0·32-1·93), p=0·589 | Disproportionately high risk/rate of inpatient falls among high income group. |
| Mosen et al (2019) | Cross-sectional | United States | 26,525 | 56·0 | 73·9 (7·4) | Fall occurrences | Neighbourhood-level socioeconomic status | Within each fall group, % that are low SES. | fall group: 14·90% non-fall group: 14·40% p=NR | Disproportionately high risk/rate of fall occurrences in low neighbourhood-level socioeconomic status group. |
| Nelson et al (2010) | Longitudinal cohort | United States | 659 | 3·8 | 54·8 (12·6) | -Fall occurrences -Injurious falls | -Education -Employment | For education, within each fall group, what % are in each exposure group. For employment, % in each fall group that are employed. | Education lowest: 36·1% fallers, 34·7% injurious fallers, 39·8% non-fallers highest: 25·0% fallers, 16·8% injurious fallers, 19·4% non-fallers p=0·65 (overall p-value) Employment fallers: 36·1% injurious fallers: 26·3% non-fallers: 24·1% p=0·04 (overall p-value) | Unclear results regarding the distribution of fall occurrences and injuriosu falls by education group. Disproportionately high risk/rate of fall occurrences and injurious falls in high employment group. |
| Noh et al (2017) | Cross-sectional | Korea | 38,627 | 56·3 | NR (all 65+) | Injurious fall | -Income -Employment | Unadjusted odds ratios (95%CI, p-value) | Income (Highest vs lowest) 0·70 (0·59-0·85), p<0·001 Employment (High vs low) 0·71 (0·68-0·75), p<0·001 | Disproportionately high risk/rate of injurious falls in low income and low employment groups. |
| Ofori-Asenso et al (2021) | Cross-sectional | United States | 1,019 | 61·7 | NR (majority 45-60) | Fall occurrences | -Education -Employment | Unadjusted odds ratios (95%CI, p-value) | Education (Lowest vs highest) 1·00 (0·68-1·47), p=NS Employment (High vs low) 0·90 (0·67-1·21), p=NS | No difference in the risk/rate of fall occurrences by education group. Disproportionately high risk/rate of fall occurrences in low employment group. |
| Ou et al (2013) | Cross-sectional | Taiwan | 1,200 | 56·3 | 59·4 (NR) | Fall occurrences | Social economic status | Within each fall group, what % are low SES (stratified by sex) | Women: 72·9% fallers, 71·6% non-fallers Men: 67·3% fallers, 61·3% non-fallers p=NR | Disproportionately high risk/rate of fall occurrences in low socioeconomic status group. |
| Pantong et al (2023) | Cross-sectional | Thailand | 12,130 | 59·8 | 71·2 (8·5) | Fall occurrences | -Education -Employment | Unadjusted odds ratios (95%CI, p-value) | Education (Highest vs lowest) 0·78 (0·62-0·99), p=0·04 Employment (High vs low) 0·52 (0·46-0·58), p<0·001 | Disproportionately high risk/rate of fall occurrences in low education and employment groups. |
| Pathania et al (2018) | Cross-sectional | India | 335 | 61·0 | 75·2 (8·6) | Fall occurrences | Education | Prevalences of falls in each exposure group | lowest: 5% highest: 22% p=0·106 (overall p-value) | Disproportionately high risk/rate of fall occurrences in high education group. |
| Pereira et al (2013) | Cross-sectional | Brazil | 6751 | NR | 70·3 (7·3) | Fall occurrences | -Education -Employment | Within each exposure group, % with fall outcome. | Education lowest: 14·1% highest: 5·6% p<0·001 (overall p-value) Employment low: 10·2% high: 9·3%  p=0·166 (overall p-value) | Disproportionately high risk/rate of fall occurrences in low education and employment groups. |
| Petersen et al (2022) | Longitudinal cohort | Germany | 11,227 | 49·1 | 64·9 (11·0) | Fall occurrences | Education | Within each exposure group, % with fall outcome (stratified by sex) | Women: 3·36% low, 4·3% high Men: 2·34% low, 2·86% high p<0·001 (overall p-value) | Disproportionately high risk/rate of fall occurrences in high education groups. |
| Peterson et al (2000) | Case-control | United States | 201 | 78·4-83·3* | 79·2 (7·5), 75·3 (5·5) † | -Injurious falls (fall-related traumatic hip fracture) | -Education -Income -Employment | Within each fall group, what % are in each exposure group. | Education lowest: 2·7% fallers, 0% non-fallers highest: 31·5% fallers, 42·2% non-fallers p=NR Income lowest: 17·1% fallers, 2·2% non-fallers highest: 21·6%, 16·7% p=NR Employment lowest: 0·9% fallers, 5·6% non-fallers highest: 7·2% fallers, 2·2% non-fallers p=NR | Disproportionately high risk/rate of injurious falls in low education group. Unclear results regarding the distribution of injurious falls by income group. Disproportionately high risk/rate of injurious falls in high employment group |
| Pimentel et al (2018a) | Cross-sectional | Brazil | 23,815 | 56·4 | NR (all 60+) | Fall occurrences | Education | Prevalence of fall outcome in each exposure group (95%CI) | low: 9·5% (8·4-10·7%) high: 7·3% (6·8-8·0%) p=NR | Disproportionately high risk/rate of fall occurrences in low education group. |
| Pitchai et al (2019) | Cross-sectional | India | 2,049 | 46·2 | 69·7 (6·9) | Fall occurrences | -Education -Socioeconomic status | Within each fall group, what % are in each exposure group. | Education lowest: 25·8% fallers, 17·6% non-fallers highest: 10·1% fallers, 28% non-fallers p=0·000 (overall p-value) SES lowest: 4·3% fallers, 1·8% non-fallers highest: 12·9% fallers, 12·5% non-fallers p=0·001 (overall p-value) | Disproportionately high risk/rate of fall occurrences in low education group. Unclear results regarding the distribution of fall occurrences by socioeconomic status group. |
| Pluijm et al (2006) | Longitudinal cohort | Netherlands | 1,365 | 51·1 | 75·3 (5·4) | Recurrent falls | Education | Unadjusted odds ratio (95%CI) | (High vs low) 1·36 (1·04-1·77), p=NR | Disproportionately high risk/rate of recurrent falls in high education group. |
| Quandt et al (2006) | Cross-sectional | United States | 691 | 49·4 | 74·1 (5·4) | Fall occurrences | Income | Within each exposure group, what % non-fallers. | lowest: 61·0% non-fallers highest: 63·0% non-fallers p=0·0034 (overall p-value) | Disproportionately high risk/rate of fall occurrences in low income group. |
| Ranaweera et al (2013) | Case-control | Sri Lanka | 1,200 | 57·0 | 71·4 (6·8) | Fall occurrences | -Education -Employment | Unadjusted odds ratios (95%CI) | Education (Low vs high) 1·07 (0·72-1·59), p=NR Employment (Low vs high) 1·56 (0·77-3·18), p=NR | Disproportionately high risk/rate of fall occurrences in low education and low employment groups. |
| Reyes-Ortiz et al (2004) | Longitudinal cohort | United States | 1,391 | 57·5-69·5* | 76·4 (5·4), 77·4 (5·8) † | Fall occurrences | Education | Mean years of education (SD) in each fall group. | fall group: 4·8 years (3·8) non-fall group: 5 years (3·9) p=0·271 | Disproportionately high risk/rate of fall occurrences in low education group. |
| Reyes-Ortiz et al (2022) | Cross-sectional | Colombia | 19,004 | 56·1 | 69·3 (7·2) | -Fall occurrences -Recurrent falls | Socioeconomic status | Unadjusted odds ratios (95%CI, p-value) | (Low vs high) fall occurrences: 0·97 (0·91-1·03), p=0·352 recurrent falls: 1·11 (1·03-1·20), p=0·009 | Mixed results regarding the distribution of risk/rate of falls between different fall outcomes for socioeconomic status. |
| Rodrigues et al (2019) | Cross-sectional | Brazil | 316 | 68·4 | 73·0 (9·0) | Fall occurrences | Education | Within each fall group, what % are in each exposure group. | lowest: 23·7% fallers, 12·4% non-fallers highest: 5·7% fallers, 9·5% non-fallers p=0·042 (overall p-value) | Disproportionately high risk/rate of fall occurrences in low education group. |
| Saari et al (2007) | Longitudinal cohort | Finland | 679 | 34·8 | 75·0 (NR), 80 (NR) † | Injurious falls | Education | Unadjusted relative risk (95% CI) | Education (Low vs high) 1·16 (0·93-1·45), p=NR | Disproportionately high risk/rate of injurious falls in low education group. |
| Sanjay et al (2022) | Longitudinal cohort | India | 260 | 58·0 | 68·7 (7·5) | Fall occurrences | -Education -Employment -Standard of living | Within each exposure group, % with fall outcome. | Education low: 48·3% high: 48% p=0·9 Employment low: 51·5% high: 41·7% p=0·1 Standard of living index low: 30·8% high: 50·2% p=0·03 | Disproportionately high risk/rate of fall occurrences in low education and low employment groups. Disproportionately high risk/rate of fall occurrences in high standard of living index group. |
| Seematter-Bagnoud et al (2006) | Longitudinal cohort | Switzerland | 690 | 61·2 | 82·4 (5·1) | Non-injurious falls | -Education -Income | Within each fall group, % that have completed high school, and % that are low income | Education fallers: 54·30% non-fallers: 49·70% p=0·47 Income fallers: 24·30% non-fallers: 23·40% p=0·87 | Disproportionately high risk/rate of non-injurious falls in high education group. Disproportionately high risk/rate of non-injurious falls in low income group. |
| Sharif et al (2018) | Cross-sectional | United Arab Emirates | 370 | 69·2 | NR (all 60+) | Fall occurrences | Education | Within each exposure group, % with fall outcome. | lowest: 75·2% highest: 21·4% p<0·001 (overall p-value) | Disproportionately high risk/rate of fall occurrences in low education group. |
| Shin et al (2009) | Longitudinal cohort | Korea | 335 | 57·0 | 72·9 (6·5) | Fall occurrences | Education | Within each fall group, what % are in each exposure group. | low: 56·82% fallers, 63·41% non-fallers high: 43·18% fallers, 36·59% non-fallers p=NS | Disproportionately high risk/rate of fall occurrences in high education group. |
| Siqueira et al (2011) | Cross-sectional | Brazil | 6,616 | 59·0 | 70·9 (8·0) | Fall occurrences | Socioeconomic status | Unadjusted prevalence ratio (95%CI) | (Lowest vs highest) 1·37 (0·97-1·95) p<0·001 (overall p-value) | Disproportionately high risk/rate of fall occurrences in low socioeconomic status group. |
| Sirohi et al (2017) | Cross-sectional | India | 456 | 56·1 | 69·4 (6·7) | Fall occurrences | Socioeconomic status | Unadjusted odds ratio (95%CI, p-value) | (Lowest vs highest) 2·1 (1·3-3·4), p=0·002 | Disproportionately high risk/rate of fall occurrences in low socioeconomic status group. |
| Smith et al (2021) | Cross-sectional | China, Ghana, India, Mexico, Russia, and South Africa | 14,585 | 54·9 | 72·6 (11·5) | Injurious falls | -Education -Wealth | Within each fall group, what % are in each exposure group. | Education lowest: 79·8% fallers, 62·7% non-fallers highest: 4·3% fallers, 6·6% non-fallers p=NR (overall p-value) Wealth lowest: 22·9% fallers, 21·6% non-fallers highest: 17·0% fallers, 19·6% non-fallers p=NR (overall p-value) | Disproportionately high risk/rate of injurious falls in low education and low wealth groups. |
| Sotoudeh et al (2023) | Cross-sectional | Iran | 653 | 50·8 | 74·1 (6·4) | Fall occurrences | -Education -Income -Housing -Worries about living expenses | Within each exposure group, % with fall outcome. | Education lowest: 48·1% highest: 25·5% p=0·01 (overall p-value) Income low: 40·1% high: 39·0% p=NS (overall p-value) Housing lowest: 44·2% highest: 38·8% p=N·S (overall p-value) Worries about living expenses low: 53·9% high: 39·8% p=0·007 (overall p-value) | Disproportionately high risk/rate of fall occurrences in low education, low income, low housing, and low worries about living expenses groups. |
| Sotoudeh et al (2018) | Cross-sectional | Iran | 653 | 50·8 | 74·1 (6·4) | Fall occurrences | -Education -Income -Housing -Worries about living expenses | Within each exposure group, % non-fallers. | Education lowest: 51·9% non-fallers highest: 74·5% non-fallers p=0·025 (overall p-value) Income low: 59·6% non-fallers high: 60·6% non-fallers p=NS (overall p-value) Housing lowest: 55·8% non-fallers highest: 61·2% non-fallers p=N·S (overall p-value) Worries about living expenses lowest: 46·1% non-fallers highest: 61·7% non-fallers p=0·030 (overall p-value) | Disproportionately high risk/rate of fall occurrences in low education, low income, low housing, and low worries about living expenses groups. |
| Souza et al (2019) | Longitudinal cohort | Brazil | 345 | 65·2 | NR (all 60+) | -Fall occurrences -Recurrent falls | Education | Unadjusted odds ratio (95%CI, p-value) | (Continuous) fall occurrences: 0·98 (0·91-1·05), p=0·535 recurrent falls: 0·94 (0·87-1·01), p=0·104 | Disproportionately high risk/rate of fall occurrences and recurrent falls in low education group. |
| Srivastava et al (2022) | Cross-sectional | India | 9,174 | 52·7 | NR (all 60+) | Injurious falls | -Education -Employment -Wealth | Within each exposure category, % with fall outcome. | Education lowest: 3·4% highest: 2·9% p=NS (overall p-value) Employment lowest: 3·8% highest: 3·9% p<0·05 (overall p-value) Wealth lowest: 3·2% highest: 2·8% p<0·05 (overall p-value) | Disproportionately high risk/rate of injurious falls in low education and low wealth groups. Disproportionately high risk/rate of injurious falls in high employment group. |
| Stolt et al (2020) | Cross-sectional | Brazil | 1,754 | 100·0 | 47·7 (8·0), 51·9 (8·8) † | Fall occurrences | Education | Unadjusted odds ratios (95%CI, p-value) (stratified by year) | (Highest vs lowest) 2007: 3·95 (1·34-11·70), p=0·013 2014:1·02 (0·48-2·16), p=0·958 | Disproportionately high risk/rate of fall occurrences in high education group. |
| Subramanian et al (2020) | Cross-sectional | India | 160 | 26·3 | 74·5 (8·9) | Fall occurrences | -Education -Socioeconomic status | Within each fall group, what % are in each exposure group. | Education low: 18·42% fallers, 14·75% non-fallers high: 81·58% fallers, 85·25% non-fallers p=0·3 (overall p-value) SES lowest: 23·68% fallers, 11·47% non-fallers highest: 23·68% fallers, 30·33% non-fallers p=0·17 (overall p-value) | Disproportionately high risk/rate of fall occurrences in low education and socioeconomic status groups. |
| Swed et al (2023) | Cross-sectional | United States | 7,132 | 51·1 | 65·82 (9·4), 64·6 (9·2) † | Injurious falls (fall-related fracture) | -Education -Income | Proportion of total sample | Education lowest: 1·2% fallers, 0·2% non-fallers highest: 2·8% fallers, 20·5% non-fallers p=NR Income low: 8·1% fallers, 47·2% non-fallers high: 5·9% fallers, 38·9% non-fallers p=NR | Unclear results regarding the distribution of injurious falls by education and income groups |
| Sylliaas et al (2012) | Longitudinal cohort | Norway | 1,147 | 73·1 | 85·0 (NR) | Fall occurrences | Education | Unadjusted relative risk (95% CI, p-value) | (Low vs high) 1·13 (1·03-1·60), p=0·027 | Disproportionately high risk/rate of fall occurrences in low education group. |
| Thakkar et al (2022) | Cross-sectional | India | 28,285 | 51·1 | NR (all 60+) | -Fall occurrences -Recurrent falls -Injurious falls | -Education -Monthly per capita consumption expenditure | Prevalence of fall outcome in each exposure category | Education lowest: 13·16% fall occurrences, 4·91% recurrent falls, 6·47% injurious falls highest: 10·63% fall occurrences, 4·55% recurrent falls, 4·18% injurious falls all fall outcome p-values <0·001 (overall p-values) Monthly per-capita expenditure lowest: 11·5% fall occurrences, 4·44% recurrent falls 5·38% injurious falls highest: 13·67% fall occurrences, 7·24% recurrent falls, 5·62% injurious falls overall p-values: 0·031 fall occurrences, 0·031 recurrent falls, 0·741 injurious falls· | Disproportionately high risk/rate of fall occurrences, recurrent falls, and injurious falls in low education group. Disproportionately high risk/rate of fall occurrences, recurrent falls, and injurious falls in high monthly per-capita expenditure group. |
| To et al (2022) | Cross-sectional | Vietnam | 340 | 62·4 | 65·7 (8·0) | Fall occurrences | -Education -Income | Unadjusted prevalence ratios (95% CI, p-value) | Education (Lowest vs highest) 1·3 (0·6-2·8), p=NS Income (Lowest vs highest) 4·1 (1·8-9·4,) p<0·001 | Disproportionately high risk/rate of fall occurrences in low education and low income groups. |
| Vieira et al (2018) | Cross-sectional | Brazil | 1,448 | 63·0 | NR (all 60+) | Fall occurrences | -Education -Employment -Economic class | Unadjusted prevalence ratios (95% CI, p-value) | Education (Lowest vs highest) 1·70 (1·22-2·39) p=0·001 (overall p-value) Employment (Low vs high) 1·60 (1·23-2·09) p=0·001 (overall p-value) Economic class (Lowest vs highest) 1·62 (1·22-2·14) p=0·001 (overall p-value) | Disproportionately high risk/rate of fall occurrences in low education, low employment, and low economic class groups. |
| Wang et al (2022) | Cross-sectional | China | 2,994 | 55·9 | NR (all 60+) | Fall occurrences | -Education -Income | Within each exposure group, % with fall outcome. | Education lowest: 10·2% highest: 10·1% p=NR Income lowest: 8·9% highest: 8·1% p=NR | Disproportionately high risk/rate of fall occurrences in low education and low income groups. |
| Wang et al (2021) | Longitudinal cohort | Australia | 273 | 100·0 | 55·2 (5·1) | Fall occurrences | -Education -Employment | Within each fall group, what % are in each exposure group. | Education lowest: 21% fallers, 33% non-fallers highest: 57% fallers, 45% non-fallers p=0·07 (overall p-value) Employment  lowest: 22% fallers· 17% non-fallers highest: 67% fallers, 61% non-fallers p=0·046 (overall p-value) | Disproportionately high risk/rate of fall occurrences in high education group. Unclear results regarding the distribution of risk/rate of fall occurrences by employment group. |
| Wang et al (2023) | Cross-sectional | China | 1,075 | 48·6 | NR (all 60+) | Fall occurrences | -Education -Income | Unadjusted odds ratios (95%CI, p-value) | Education (Lowest vs highest) 5·93 (2·89-12·15), p<0·01 Income (Lowest vs highest) 4·19 (1·77-9·96), p<0·01 | Disproportionately high risk/rate of fall occurrences in low education and low income groups. |
| Wen et al (2021) | Longitudinal cohort | China | 1,238 | 49·0-56·9* | 59·4 (9·3), 61·8 (9·0) † | Fall occurrences | Education | Within each fall group, what % are in each exposure group. | lowest: 33·2% fallers, 24·9% non-fallers highest: 28·3% fallers, 34·6% non-fallers p=0·02 (overall p-value) | Disproportionately high risk/rate of fall occurrences in low education group. |
| Wojszel et al (2004) | Cross-sectional | Poland | 456 | 61·8-64·2* | 79·8 (5·0), 80·8 (4·8) † | Fall occurrences | Scale of living standards | Within each exposure category, % non-fallers. | lowest: 30·4% non-fallers highest: 60·2% non-fallers p=0·006 (overall p-value) | Disproportionately high risk/rate of fall occurrences in low scale of living standards group. |
| Woo et al (2009) | Longitudinal cohort | Hong Kong | 4,000 | 50·1 | NR (all 65+) | Recurrent falls | Education | Unadjusted odds ratio (95%CI) (stratified by sex) | (Highest vs lowest) males: 1·23 (0·78-1·96) females: 1·77 (1·09-2·88) p=NR | Disproportionately high risk/rate of recurrent falls in high education group. |
| Yeh et al (2017) | Longitudinal cohort | Taiwan | 171 | 66·7 | 78·1 (NR) | Fall occurrences | Education | Within each exposure group, % non-fallers. | lowest: 66·15% non-fallers highest: 88·89% non-fallers p=0·267 (overall p-value) | Disproportionately high risk/rate of fall occurrences in low education group. |
| Zhao et al (2018) | Cross-sectional | United States | 5,930 | 57·2 | NR (all 65+) | Fall occurrences | Education | Within each fall group, % in each exposure group. | lowest: 28·8% fallers, 22·1% non-fallers highest: 44·3% fallers, 49·4% non-fallers p<0·001 | Disproportionately high risk/rate of fall occurrences in low education group. |
| Zhou et al (2022) | Longitudinal cohort | China | 5,110 | 45·3 | 67·3 (5·9) | Fall occurrences | Education | Within each exposure category, % with fall outcome. | lowest: 22·23% highest: 17·56% p=0·028 (overall p-value) | Disproportionately high risk/rate of fall occurrences in low education group. |
| Zimba Kalula et al (2015) | Cross-sectional | South Africa | 837 | 76·5 | 74·0 (6·4) | -Fall occurrences -Recurrent falls | -Education -Occupation | Unadjusted odds ratios (95%CI, p-value) (stratified by baseline, follow-up, and baseline and follow-up) | Education (Highest vs lowest) Fall occurrences: baseline: 1·64 (0·67-3·98), p=NS follow-up: 0·84 (0·27-2·56), p=NS baseline and follow-up: 0·56 (0·14-2·18), p=NS Recurrent falls: baseline: 0·80 (0·20-3·24), p=NS follow-up: 0·42 (0·09-1·94), p=NS Occupation (Highest vs lowest) Fall occurrences: baseline: 3·73 (2·18-6·36), p<0·001 follow-up: 1·83 (1·01-3·32), p<0·001 baseline and follow-up: 3·41 (1·46-7·95), p<0·001 Recurrent falls: baseline: 2·72 (1·29-5·71), p<0·05 follow-up: 2·94 (1·13-7·61), p<0·05 | Disproportionately high risk/rate of fall occurrences and recurrent falls in high occupation group. Mixed results regarding the distribution of fall occurrences and recurrent falls by education group. |
| Zonzini Gaino et al (2019) | Cross-sectional | Brazil | 113 | 86·7 | 54·7 (NR), 58·6 (NR), 55·8 (NR)† | -Fall occurrences -Recurrent falls | -Education -Income | Within each fall group, mean (SD) years of formal education and mean (SD) monthly income in Brazilian Reals. | Education sporadic fallers: 6·5 (3·6) recurrent fallers: 6·9 (4·3) non-fallers: 8·0 (3·6) p=0·1837 (overall p-value) Income sporadic fallers: 1453·1 (743·3) recurrent fallers: 1277·0 (588·9) non-fallers: 1676·8 (925·9) p=0·1665 (overall p-value) | Disproportionately high risk/rate of fall occurrences and recurrent falls in low education and low income groups. |

* range of % female across multiple strata where overall % not reported

† mean (SD) age across multiple strata where overall mean (SD) not reported

Abbreviations: SD, standard deviation: IQR, interquartile range; CI, confidence interval; NR, not reported: NA, not applicable; NS, not significant

### Specific considerations

Number of SEP indicators:

A thorough comparison of the measurement of SEP indicators was undertaken between studies (Tables S3 and S5), which showed that very few studies on this topic cover a wide range of different SEP indicators – with most only covering one indicator. Among the 14 studies explicitly aiming to study SEP and falls, only five included multiple indicators of SEP within adjusted results. Of these, only three contained adjusted results on three or more SEP indicators (Table S3). Among the nine other studies with relevant adjusted results, only one included multiple indicators of SEP (Table S5).

Overadjustment of mediators:

In our main synthesis, we only report the results of the most appropriate model within a given study. Yet, of the results synthesised from the most appropriate model for the 14 studies explicitly aiming to study SEP and falls, seven still overadjusted for variables that are likely mediators of the relationship between SEP and falls (e.g. chronic conditions, acute conditions, body mass index, depression, and smoking). Further, seven of these 14 studies also contained results from other adjusted models or from more complex analysis (Table S9). These additional results show further issues with overadjustment of mediators within these studies, with four of these seven studies having overadjusted for likely mediators within these additional results.

### Table S10 – Risk of bias assessment summary

|  | **Author & Date** | **1. Study Participation** | **2. Study Attrition** | **3. Exposure Measurement** | **4. Outcome measurement** | **5. Study Confounding** | **6. Statistical Analysis** | **7. Overadjustment Bias** |
| --- | --- | --- | --- | --- | --- | --- | --- | --- |
| **Studies explicitly aiming to study SEP and falls** | Brown et al (2023) | Moderate | N/A | Low | High | Low | Low | High |
|  | Gauchard et al (2006) | High | N/A | Low | Moderate | Moderate | Low | Low |
|  | Gribbin et al (2009) | Low | High | Low | Moderate | Low | Low | Low |
|  | Hong et al (2020) | High | N/A | Low | High | Low | Low | High |
|  | Jyvakorpi et al (2019) | High | N/A | Low | High | High | High | N/A |
|  | Khalatbari-Soltani et al (2021) | Moderate | Low | Low | Moderate | Low | Low | Low |
|  | Kiadaliri et al (2018) | Moderate | Moderate | Low | Low | Low | Low | Low |
|  | Li et al (2014) | Moderate | High | Low | Low | Low | Low | High |
|  | Liu and Hu (2022) | High | High | Moderate | High | Low | Low | Low |
|  | Lo et al (2016) | Moderate | Moderate | Low | High | Low | Low | High |
|  | Ryu et al (2017) | Low | Low | Low | Low | Low | Low | Moderate |
|  | Sairafian et al (2019) | Moderate | N/A | Low | High | Low | Low | High |
|  | Syddall et al (2009) | Moderate | N/A | Low | High | Moderate | Moderate | High |
|  | Trujillo et al. (2011) | Moderate | N/A | Low | High | Moderate | Low | High |
| **Other studies with relevant adjusted models** | Ho et al (1996) | Low | N/A | High | Moderate | Moderate | Low | High |
|  | Kelsey et al (2010) | Moderate | Moderate | Low | Low | Moderate | Low | Low |
|  | Nordstrom et al (1996) | Low | N/A | Moderate | Low | High | Low | Low |
|  | Pimentel et al (2018b) | Low | N/A | Moderate | Moderate | Moderate | Low | Low |
|  | Rodrigues et al (2014) | Low | N/A | Low | High | Moderate | Low | Low |
|  | Salva et al (2004) | Low | High | Moderate | Low | Moderate | Low | Low |
|  | Seo et al (2022) | Moderate | Moderate | Moderate | Moderate | Low | Moderate | Moderate |
|  | Sharma et al (2018) | Low | Moderate | Low | Moderate | Low | Low | High |
|  | Sprince et al (2003) | Moderate | N/A | Moderate | Moderate | High | Low | Low |
|  | Total high risk | 4 | 4 | 1 | 9 | 3 | 1 | 9 |
|  | Total moderate risk | 11 | 5 | 6 | 8 | 8 | 2 | 2 |
|  | Total low risk | 8 | 2 | 16 | 6 | 12 | 20 | 11 |

Abbreviations: NA = not applicable

* study attrition not applicable to these studies given their study types (cross-sectional or case-control)

† overadjustment bias not applicable as only unadjusted results were reported

### Synthesis of risk of bias assessment results

Overall

With regards to bias in study participation, out of 23 studies with either explicit aims to study SEP and falls or with relevant adjusted models, 11 were found to have a moderate risk of bias, eight with low risk of bias, and only four with high risk of bias (Table S10). The study attrition item only applied to 11 of the 23 studies, as the remaining 12 were all either cross-sectional or case-control studies. Of these, four were at high risk of study attrition bias, five at moderate risk, and two at low risk. Generally, risk of bias for exposure measurement was low (n =16), with only one study having a high risk of exposure measurement bias, and six having moderate risk. A high risk of bias was common for outcome measurement (n=9), with another eight having moderate risk, and six having low risk. For study confounding, bias was generally low (n=12) or moderate (n=8), with only three studies having a high risk of bias. Similarly, bias in the statistical analysis was rare, with 20 studies having a low risk of bias, two a moderate risk, and only one a high risk. Finally, the item for overadjustment bias applied to 22 studies as the remaining study only reported unadjusted results. Of these, a high risk of overadjustment bias was common (n=9), while 11 studies had a low risk of bias, and two had a moderate risk of bias.

Comparison between the 14 studies explicitly aiming to study SEP and falls and the nine other studies with relevant adjusted results

There were notable differences in risk of bias between the 14 studies explicitly aiming to study SEP and falls and the nine other studies with relevant adjusted models. Namely, risk of study participation, outcome measurement, and overadjustment biases were generally higher among the 14 studies explicitly aiming to study this topic compared to the nine other studies with relevant adjusted models. Meanwhile, risk of exposure measurement and study confounding biases were generally lower among the 14 studies explicitly aiming to study this topic compared to the nine other studies with relevant adjusted models.

Comparison in risk of bias between studies according to results

Of the 23 studies represented in our main synthesis, four reported estimates of effect indicating that higher SEP is adversely associated with risk/rate of falls. Of these four studies, high risk was evident in two for overadjustment bias, one for confounding bias, one for outcome measurement bias, and one for exposure measurement bias. Further, three of these studies had a moderate risk of study participation bias, one of exposure measurement bias, two of outcome measurement bias, two of study confounding bias, and one of study attrition bias.

Comparatively, 14 of the 23 studies reported estimates of effect indicating that higher SEP is protectively associated with risk/rate of falls. Among these, two had a high risk of study participation bias, three of study attrition bias, four of outcome measurement bias, one of study confounding bias, and six of overadjustment bias. Additionally, six had a moderate risk of study participation bias, four of study attrition bias, four of exposure measurement bias, five of outcome measurement bias, five of study confounding bias, two of statistical analysis bias, and two of overadjustment bias.

### Table S11 – Association between socioeconomic indicators and falls outcome from studies with explicit aim – results from various level of adjustment models

| **Author** | **Additional Model(s)** | **Additional Model(s)** | **Additional Model(s)** | **Additional Model(s)** | **Additional Model(s)** | **Additional Model(s)** |
| --- | --- | --- | --- | --- | --- | --- |
| Brown et al (2023) | Income (stratified by race)  Non-Hispanic White: IRR (continuous) 1·02 (0·99-1·05), p = NR  Non-Hispanic Black: IRR (continuous) 1·05 (0·99-1·11), p = NR  (adjusted for sex, race/ethnicity, age, educational attainment, body mass index, depression score, anaemic status, number of chronic conditions, density of neighbourhood clinical-care facilities, density of neighbourhood pharmacies, health insurance status, and neighbourhood quartile of persons living in poverty) | Income (segmented regression stratified by age)  45-64 y/o – breakpoint: 13,826 USD (4,666-22,986)  - Below breakpoint: IRR (continuous) 0·34 (0·08-1·36), p = NR  - Above breakpoint: IRR (continuous) 0·95 (0·87-1·05), p = NR  65+ y/o – breakpoint: 20,557 USD (3,456-37,656)  - Below breakpoint: IRR (continuous) 1·09 (0·23-1·82), p = NR  - Above breakpoint: IRR (continuous) 0·64 (0·99-1·21), p = NR  (adjusted for sex, race/ethnicity, age, educational attainment, body mass index, depression score, anaemic status, number of chronic conditions, density of neighbourhood clinical-care facilities, density of neighbourhood pharmacies, health insurance status, and neighbourhood quartile of persons living in poverty) | Income (segmented regression stratified by race)  Non-Hispanic White – breakpoint: 15,914 USD (4,315-27,514)  - Below breakpoint: IRR (continuous) 0·45 (0·11-1·74), p = NR  - Above breakpoint: IRR (continuous) 1·04 (0·97-1·21), p = NR  Non-Hispanic Black – breakpoint: 48,058 USD (32,841-63,276)  - Below breakpoint: IRR (continuous) 0·56 (0·43-0·74), p = NR  - Above breakpoint: IRR (continuous) 1·37 (1·05-1·79), p = NR  (adjusted for sex, race/ethnicity, age, educational attainment, body mass index, depression score, anaemic status, number of chronic conditions, density of neighbourhood clinical-care facilities, density of neighbourhood pharmacies, health insurance status, and neighbourhood quartile of persons living in poverty) | .. | .. | .. |
| Khalatbari-Soltani et al (2021) | Education  IRR (low vs high) 0·88 (0·66-1·77), p = NR  (adjusted for age)  Education (stratified by country of birth)  Australian-born: IRR (low vs high) 1·68 (1·18-2·40), p = NR  Migrant born in non-main English-speaking countries: IRR (low vs high) 0·41 (0·24-0·71), p = NR  (adjusted for age)  Education (stratified by country of birth)  Australian-born: IRR (low vs high) 1·47 (1·10-1·96), p = NR  Migrant born in non-main English-speaking countries: IRR (low vs high) 1·47 (1·10-1·96), p = NR  (adjusted for age and living arrangement) | Occupation  IRR (low vs high) 1·06 (0·85-1·32), p = NR  (adjusted for age)  Occupation (stratified by country of birth)  Australian-born: IRR (low vs high) 1·47 (1·10-1·96), p = NR  Migrant born in non-main English-speaking countries: IRR (low vs high) 1·47 (1·10-1·96), p = NR  (adjusted for age)  Occupation (stratified by country of birth)  Australian-born: IRR (low vs high) 1·45 (1·09-1·93), p = NR  Migrant born in non-main English-speaking countries: IRR (low vs high) 1·45 (1·09-1·93), p = NR  (adjusted for age and living arrangement) | Income  IRR (low vs high) 1·01 (0·83-1·23), p = NR  (adjusted for age)  Income (stratified by country of birth)  Australian-born: IRR (low vs high) 1·18 (0·89-1·57), p = NR  Migrant born in non-main English-speaking countries: IRR (low vs high) 1·18 (0·89-1·57), p = NR  (adjusted for age)  Income (stratified by country of birth)  Australian-born: IRR (low vs high) 1·12 (0·84-1·50), p = NR  Migrant born in non-main English-speaking countries: IRR (low vs high) 1·12 (0·84-1·50), p = NR  (adjusted for age and living arrangement) | Housing  IRR (low vs high) 1·09 (0·82-1·45), p = NR  (adjusted for age)  Housing (stratified by country of birth)  Australian-born: IRR (low vs high) 1·11 (0·76-1·61), p = NR  Migrant born in non-main English-speaking countries: IRR (low vs high) 1·11 (0·76-1·61), p = NR  (adjusted for age)  Housing (stratified by country of birth)  Australian-born: IRR (low vs high) 1·01 (0·69-1·48), p = NR  Migrant born in non-main English-speaking countries: IRR (low vs high) 1·01 (0·69-1·48), p = NR  (adjusted for age and living arrangement) | Cumulative socioeconomic status  IRR (low vs high) 0·97 (0·79-1·20), p = NR  Cumulative socioeconomic status  (stratified by country of birth)  Australian-born: IRR (low vs high) 1·37 (1·00-1·87), p = NR  Migrant born in non-main English-speaking countries: IRR (low vs high) 0·83 (0·58-1·18), p = NR  (adjusted for age)  Cumulative socioeconomic status  (stratified by country of birth)  Australian-born: IRR (low vs high) 1·31 (0·96-1·79), p = NR  Migrant born in non-main English-speaking countries: IRR (low vs high) 0·82 (0·58-1·16), p = NR  (adjusted for age and living arrangement) | Cumulative socioeconomic status without education  IRR (low vs high) 1·06 (0·82-1·36), p = NR  Cumulative socioeconomic status without education (stratified by country of birth)  Australian-born: IRR (low vs high) 1·47 (1·00-2·19), p = NR  Migrant born in non-main English-speaking countries: IRR (low vs high) 1·47 (1·00-2·18), p = NR  (adjusted for age)  Cumulative socioeconomic status without education (stratified by country of birth)  Australian-born: IRR (low vs high) 1·38 (0·93-2·05), p = NR  Migrant born in non-main English-speaking countries: IRR (low vs high) 1·38 (0·93-2·05), p = NR  (adjusted for age and living arrangement) |
| Kiadaliri et al (2018) | Education (stratified by sex)  Women: SII 1·6 (-2·8-5·9)  Men: SII 15·5 (9·8-21·3)  (adjusted for marital status and place of birth) | Education (stratified by sex)  Women: RII 1·2 (0·8-1·8)  Men: RII 2·2 (1·6-3·0)  (adjusted for marital status and place of birth) | Education (stratified by sex)  Women: PAF (%) 6·0 (-21·3-27·1)  Men: PAF (%) 33·7 (18·6-46·0)  (adjusted for marital status and place of birth) | ·· | ·· | ·· |
| Liu and Hu (2022) | Education (stratified by region)  Urban: β (low vs high) -0·0073 (NR), p = NS  Outskirts: β (low vs high) -0·3947 (NR), p = NS  Rural: β (low vs high) 0·0326 (NR), p = NS  (adjusted for gender, age, home address, residence category, and spouse) | Income (stratified by region)  Urban: β (high vs low) -0·3828 (NR), p < 0·1  Outskirts: β (high vs low) -0·6417 (NR), p = NS  Rural: β (high vs low) -0·2341 (NR), p < 0·01  (adjusted for gender, age, home address, residence category, and spouse) | Occupation (stratified by region)  Urban: β (high vs low) 0·5616 (NR), p < 0·1  Outskirts: β (high vs low) -0·0066 (NR), p < 0·01  Rural: β (high vs low) -0·0118 (NR), p < 0·01  (adjusted for gender, age, home address, residence category, and spouse) | Socioeconomic status (mediation analysis)  Direct effect: β (continuous) -0·0052 (NR), p = NS  Indirect effect (through health shock): β (continuous) -0·0066 (NR), p < 0·01  Total effect: β (continuous) -0·0118 (NR), p < 0·01 | ·· | ·· |
| Ryu et al (2017) | Socioeconomic status  HR (highest vs lowest) 0·64 (0·48-0·86)  (adjusted for age, sex, body mass index, education, and number of chronic conditions) | Socioeconomic status  HR (highest vs lowest) 0·55 (0·44-0·70)  (adjusted for age, sex, body mass index, education, and number of chronic conditions) | Socioeconomic status  HR (highest vs lowest) 0·58 (0·44-0·76)  (adjusted for age, sex, and body mass index) | ·· | ·· | ·· |
| Syddall et al (2009) | Social class in adulthood (among women)  OR (lowest vs middle) 1·1 (1·0-1·2), p = 0·07  (adjusted for age, height, weight adjusted for height, smoking, alcohol, and marital status) | Social class in adulthood (among women)  OR (lowest vs middle) 1·1 (1·0-1·2), p = 0·06  (adjusted for age, height, weight adjusted for height, smoking, alcohol, marital status, and walking speed) | Social class in adulthood (among women)  OR (lowest vs middle) 1·1 (1·0-1·2), p = 0·05  (adjusted for age, height, weight adjusted for height, smoking, alcohol, marital status, walking speed, ischemic heart disease, and hypertension) | ·· | ·· | ·· |
| Trujillo et al (2011) | Income  Chile: β (continuous) -0·016 (NR), p = NS  Argentina: β (continuous) -0·0080 (NR), p = NS  Mexico: β (continuous) -0·025 (NR), p = NS  (adjusted for age, sex, school, home ownership, and enough resources to cover daily needs) | Income  Chile: β (continuous) -0·103 (NR), p < 0·05  Argentina: β (continuous) -0·010 (NR), p = NS  Mexico: β (continuous) -0·028 (NR), p = NS  age, sex, total children alive, marital status, school, home ownership, enough resources to cover daily needs, number of household members, height, weight, living in rural areas first 5 years of life, serious health problems first 15 years of life, starvation in the first 15 years of life, hypertension, diabetes, cancer, chronic lung disease, heart disease, stroke, arthritis, rheumatism or osteoarthritis, incontinence, memory, cognitive scores, emotional, nervous, or psychiatric problem, self-rated health status, activities of daily living index, and independent activities of daily living index) | ·· | ·· | ·· | ·· |

Abbreviations: NR, not reported; NS, not significant; USD, United States Dollar; IRR, incidence rate ratio; SII, slope index of inequality; RII, relative index of inequality; PAF, population attributable fraction; β, beta coefficient; HR, hazard ratio; OR, odds ratio
